# Clinical dimensions along the progressive nonfluent variant primary progressive aphasia spectrum

**DOI:** 10.1101/2023.04.18.23288702

**Authors:** Ignacio Illán-Gala, Diego L. Lorca-Puls, Zoe Ezzes, Jessica Deleon, Zachary A. Miller, Sara Rubio-Guerra, Miguel Santos-Santos, David Gómez-Andrés, Lea T. Grinberg, Salvatore Spina, Joel H. Kramer, Lisa Wauters, Maya L. Henry, Bruce L. Miller, William W. Seeley, Maria Luisa Mandelli, Maria Luisa Gorno-Tempini

## Abstract

It is debated whether primary progressive apraxia of speech (PPAOS) and progressive agrammatic aphasia (PAA) belong to the same clinical spectrum traditionally termed nonfluent/agrammatic variant primary progressive aphasia (nfvPPA) or exist as two completely distinct syndromic entities with specific pathologic/prognostic correlates. We analyzed speech, language, and disease severity features in a comprehensive cohort of patients with a progressive motor speech impairment and/or agrammatism to ascertain any evidence of the existence of naturally occurring, non-overlapping syndromic entities (e.g., PPAOS and PAA) in our data. We also assessed if data-driven latent clinical dimensions with etiologic/prognostic value could be identified. We included 98 participants with progressive motor speech impairment and/or agrammatism, with 43 having an autopsy-confirmed neuropathological diagnosis. Speech pathologists assessed motor speech features indicative of dysarthria and apraxia of speech (AOS). Quantitative expressive/receptive agrammatism measures were obtained and compared with healthy controls. Baseline and longitudinal disease severity was evaluated using the Clinical Dementia Rating sum-of-boxes (CDR-SB). We investigated the data’s clustering tendency to form robust symptom clusters and employed principal component analysis to extract data-driven latent clinical dimensions (LCD). The longitudinal CDR-SB change was estimated utilizing linear mixed-effects models. Of the participants included in this study, 91 conformed to previously reported clinical profiles (69 with AOS and agrammatism, 18 PPAOS, and 4 PAA). The remaining seven participants were characterized by nonfluent speech and dysarthria without apraxia of speech or agrammatism. No baseline clinical features differentiated between FTLD neuropathological subgroups. Critically, the Hopkins statistic dismissed the presence of non-overlapping syndromic clusters in the entire sample (.45 with values near 0.5 indicating random data). Three data-driven components accounted for 71% of the variance ([i] severity-agrammatism, [ii] prominent AOS, and [iii] prominent dysarthria). The component typified by prominent dysarthria was more specific to patients with Progressive Supranuclear Palsy (4/5 [80%] participants with autopsy in this group had PSP), while the severity-agrammatism component predicted a faster CDR-SB increase. Higher dysarthria severity, reduced words per minute, and expressive and receptive agrammatism severity at baseline independently predicted accelerated disease progression (as measured by the CDR-SB score). Our findings indicate that PPAOS and PAA, rather than exist as completely distinct syndromic entities, constitute a clinical continuum strongly predictive of underlying Frontotemporal Lobar Degeneration (FTLD, 66% 4R tauopathy, 16% Pick’s disease, 9.3% FTLD TDP type A, and 12% other pathologies). While highlighting the graded distinctions (rather than sharp boundaries) that characterize the nfvPPA spectrum may be useful for establishing early clinical rehabilitation strategies, novel clinical and biological markers are needed to improve clinical-pathological correlations.

## Introduction

The non-fluent/agrammatic variant of Primary Progressive Aphasia (nfvPPA) is a neurodegenerative clinical syndrome currently defined by effortful speech (mainly caused by apraxia of speech [AOS]), and varying degrees of expressive agrammatism.^1^ Effortful speech and expressive agrammatism in the setting of relatively spared single-word comprehension and object knowledge represent the two core features of nfvPPA and can appear combined or in isolation.^2^ But, in addition to AOS and agrammatic production errors, a wide range of changes in motor speech and grammar abilities can be noted in nfvPPA, and these features frequently overlap with other cognitive, speech, and language deficits like dysarthria or executive dysfunction.^3^ Importantly, previous studies have documented the existence of distinguishable speech-language profiles in patients with nfvPPA arising from the differential expression of AOS versus expressive agrammatism.^4–7^ For example, the term “progressive agrammatic aphasia” or “agrammatic variant PPA”^5, 8^ has been used to refer to patients with expressive agrammatism as the most prominent presenting symptom in the absence of AOS. In contrast, patients exhibiting AOS as the most salient clinical feature without clear concomitant aphasia (including expressive agrammatism) have been ascribed the term “primary progressive apraxia of speech” (PPAOS).^6^ Some authors suggest that PPAOS and PAA should be considered separate syndromic entities because they may have specific etiologic and prognostic features. However, existing evidence does not clearly indicate whether these phenotypes represent a clinical continuum or exist as distinct syndromic entities with specific anatomical, pathological, and prognostic correlates. It remains unclear whether syndromic definitions or data-driven approaches will significantly improve clinical-pathological correlations and disease progression.^9^ Finally, to date, the most successful speech therapy approach to nfvPPA rehabilitation,^10^ targets both motor speech and agrammatic symptoms. This approach is justified by evidence from longitudinal studies showing that, even in cases where AOS and agrammatism are initially isolated, they eventually co-occur as the disease progresses. From a pathological perspective, large autopsy-proven studies have shown that patients presenting with both AOS and agrammatism, PPAOS, and PAA are typically associated with Progressive Supranuclear Palsy (PSP) and Corticobasal Degeneration (CBD).^11^ But, less frequently, these phenotypes are also the clinical presentation of Pick’s disease,^12^ or other Frontotemporal Lobar Degeneration (FTLD) subtypes, like the phosphorylated 43-kDa TAR DNA-binding protein inclusions (FTLD-TDP) subtypes.^13^ Current syndromic classifications have proven helpful in discriminating between patients with FTLD versus Alzheimer’s disease (AD),^2, 14, 15^ but have failed to predict FTLD subtypes (i.e., PSP vs CBD) or longitudinal decline at the single-subject level.^11^ Refining the classification of patients within the nfvPPA-spectrum (nfvPPA-S) could improve clinical-pathological correlations,^16^ and the prediction of progression rate, both of which are essential steps to designing successful trials targeting molecular mechanisms of pathology.^17^

Data-driven classifications allow the recognition of individual differences within phenotypes and may provide valuable insights for the refinement of diagnostic labels and the development of precision medicine approaches.^18^ Unfortunately, previous studies exploring data-driven subtypes of nfvPPA were limited by their relatively small sample size and lack of longitudinal and autopsy data.^19, 20^ In this study, we aimed to characterize the speech, language, and cognitive features among patients with a broad spectrum of nonfluent speech and/or language impairments and explore the existence of data-driven latent clinical dimensions (LCD) with etiologic and/or prognostic value.

We hypothesize that PPA and PPAOS represent a clinical continuum encompassing several endophenotypes with prognostic implications that could be useful to improve the clinical-pathological correlations.

## Material and Methods

### Participant selection

We searched the Memory and Aging Center of the University of California, San Francisco (MAC-UCSF) database to identify participants within the nfvPPA-S, as defined by a progressive motor speech impairment (either apraxia of speech or dysarthria) and/or expressive agrammatism that completed a comprehensive assessment of speech-language skills.^2^ Importantly, participants in this study were not required to exhibit prominent aphasia at the time of diagnosis. Consequently, not all individuals classified within the nfvPPA-S in this investigation strictly satisfied the core diagnostic criteria for primary progressive aphasia.^2, 21^ We excluded participants at an advanced disease stage (i.e., those with a Mini-Mental State Examination (MMSE)^22^ score ≤ 10, or, alternatively, a Clinical Dementia Rating (CDR)^23^ global score ≥ 3) because the inclusion of outliers at an advanced disease stage could bias the results. We also excluded participants meeting diagnostic criteria for the logopenic or semantic variants of primary progressive aphasia, or with prominent behavioral or motor symptoms meeting diagnostic criteria for the behavioral variant of frontotemporal dementia, PSP or CBD.^24, 25^ The diagnosis of all participants was imaging-supported, and vascular disease was ruled out. Ninety-eight participants recruited at MAC-UCSF between September 1999 and January 2021 were included in the study. In participants with multiple clinical assessments, we selected the first visit with a complete speech-language assessment because clinical features of FTLD-related syndromes tend to converge with disease progression.^3^

### Genetic and neuropathological assessment

A neuropathological evaluation was available in a subgroup of the participants (n=43). Neuropathological diagnoses and genetic analyses were performed as previously described.^26, 27^ Briefly, participants were classified post-mortem into FTLD major molecular classes and subtypes, and genetic screening was conducted for mutations known to cause autosomal dominant FTLD or AD in the participants giving their informed consent.^28^

## Clinical assessment

### General measures of cognitive and functional impairment

All participants underwent a complete clinical history, physical examination, and neuropsychological evaluation. We used a previously reported battery of neuropsychological tests to assess major cognitive domains. In addition, all participants underwent a comprehensive standardized speech and language assessment, as previously described. ^29–31^ The CDR (available for 95 [97%] of the participants at baseline), and the CDR Staging Instrument PLUS National Alzheimer’s Coordinating Center Behavior and Language Domains (CDR plus NACC FTLD, available for 84 [86%] of the participants at baseline) were adopted as general measures of disease severity.^23, 32^ The MMSE was also recorded at baseline and used as a general measure of cognitive impairment.^22^ We also report the estimated age at symptom onset, sex, years of education, and age at diagnosis. In participants with more than one visit, the longitudinal decline was characterized using the CDR sum-of-boxes (CDR-SB) when available. In longitudinal analyses, the CDR-SB was prioritized over the CDR plus NACC FTLD sum-of-boxes (CDR plus NACC FTLD-SB) because the former resulted in fewer missing values.

### Assessing apraxia of speech and dysarthria through a structured motor speech evaluation

To elicit a wide range of motor speech behaviors, the participant was asked, as part of our motor speech evaluation (MSE),^33^ to complete a collection of tasks such as vowel prolongation, alternating motion rate, sequential motion rate, multiple repetitions of monosyllabic words, multiple repetitions of multisyllabic words, repetition of words of increasing length, and the reading of a brief, phonetically balanced paragraph. Based on the observed motor speech ability of the patient, a certified speech-language pathologist assigned a clinical severity rating for AOS and separately for dysarthria on a scale from 0 (within normal limits) to 7 (profound). A list of deviant motor speech characteristics used to perceptually judge the presence and severity of AOS and/or dysarthria is provided in **Supplementary Table 1**.

### Assessing expressive agrammatism and speech fluency through a picture description task

To further characterize grammar processing abilities, we enriched the standard speech and language protocol with additional quantitative measures of syntactic ability and speech fluency by analyzing connected speech samples of the participants. To assess expressive agrammatism, the participant was prompted to describe a visual scene in as much detail as possible by using sentences. Audio-recorded connected speech samples describing the “picnic” scene from the Western Aphasia Battery^34^ were sent to www.saltsoftware.com for transcription, coding, and analysis. By running the coded transcripts through the Systematic Analysis of Language Transcripts (SALT) software, a set of measures was generated, of which we selected exclusively those capturing the accuracy and complexity of the sentences produced by the patient (consistent with the definition of expressive agrammatism) and the number of words per minute as a general measure of speech fluency/rate. Four SALT-derived measurements were assessed: (1) % utterances with omission and/or commission errors (%UtWErrors), a measure of morphosyntactic accuracy that captures missing function/content words, omitted bound morphemes, inappropriate word choice, and/or incorrect morphosyntactic form; (2) mean length of utterance (MLU), a measure of morphosyntactic complexity that captures mean sentence length in words; (3) subordination index (SI), a measure of morphosyntactic complexity that captures the ratio of the total number of clauses to the total number of utterances; and (4) words per minute (WPM) as a general measure of speech fluency. Only complete (not abandoned or interrupted), intelligible (without any unintelligible segments), and verbal utterances (that contained at least one verbalized word) contributed to the calculation of these quantitative, continuous measures.

### Assessing receptive agrammatism through a sentence comprehension task

To assess receptive agrammatism, the participant was prompted to complete either of two auditory sentence-to-picture matching tasks that are part of our comprehensive speech-language assessment battery. The first task involved a representative range of sentence types, varying in both length and complexity, taken from the Curtiss Yamada Comprehensive Language Evaluation (CYCLE) as previously reported.^35^ Each patient was instructed to match the meaning of an auditorily-presented sentence with the corresponding line drawing in a three-or four-picture array. The second task was loosely based on the first and has been previously described.^36^ To reduce the number of participants with missing data for receptive agrammatism, we also considered the syntax comprehension score from our bedside neuropsychological battery. This score includes five sentences from the Boston Diagnostic Aphasia Examination, ranges from 0 (worst) to 5, and was moderately correlated with the other sentence comprehension tasks (Rho=.650, *p*<.001).

### Assessing other relevant cognitive and behavioral aspects

Because speech and language impairment could impact neuropsychological performance in tests with a high verbal load, we distinguish between verbal and non-verbal tests when defining the different cognitive domains. Hence, memory was assessed with the delayed recognition of words from the California verbal learning test^37^ and the delayed recall of the Benson figure.^38^ Visuospatial ability was assessed with the number location subtest of the visual object space perception battery and the copy of the Benson figure.^38^ Verbal measures of executive dysfunction included reverse digit span and the Stroop test,^39^ while non-verbal measures included the design fluency subscale of the Delis-Kaplan Executive Function Scale and the number of correct trials in 1 minute from the modified trail-making test.^40, 41^ Naming and word comprehension were assessed with the 15-item version of the Boston naming test,^42^ and the Peabody Picture Vocabulary Test, respectively. Repetition was assessed with the Western Aphasia Battery (WAB). The caregiver-distress scores of the Neuropsychiatric Inventory (NPI) were recorded to characterize behavioral changes.^43^

### Definition of speech, language, and cognitive domains

To create a single index of expressive grammar ability, the %UtWErrors, MLU, and SI scores were combined after their normalization. Similarly, scores from the two sentence comprehension tasks included in our speech-language assessment bettery were combined with the syntax comprehension score from the bedside neuropsychological battery to create an index of receptive grammar ability. In addition, cognitive variables measuring non-overlapping neuropsychological domains were grouped based on *a priori* knowledge from the authors and their observed correlation. These domains include: (i) verbal measures of executive function (reverse digit span and the Stroop test); (ii) non-verbal measures of executive function (design fluency subscale of the Delis-Kaplan Executive Function Scale, and the number of correct trials in 1 minute at the modified trail-making test); (iii) verbal and non-verbal measures of memory (the California verbal learning test and the delayed recall of the Benson figure); and (iv) non-verbal measures of visuospatial function (the number location subtest of the visual object space perception battery and the copy of the Benton figure). The Z-scores of variables within the same domain were averaged to maximize the number of participants with at least one measurement for each clinical domain. By grouping variables taxing similar neuropsychological domains, we avoided both data imputations and the exclusion of participants with missing data in a few variables.

### Definition of impaired performance

MSE ratings for apraxia of speech or dysarthria greater than 0, as judged by a certified speech-language pathologist, were considered impaired. However, the performance of an individual patient on the picture description and sentence comprehension tasks cannot be labeled as “impaired” without reference to a normative sample of neurologically intact controls. For these tasks, we used a previously validated Bayesian method to determine if a patient’s task performance fell within the impaired range (while covarying out the effects of age and sex).^44^ SALT-derived measures of expressive agrammatism (%UtWErrors, MLU, and SI), and WPM were considered individually relative to a group of 18 neurologically intact controls (mean age ± SD = 71.69 ± 5.27 years, range = 56 – 79 years; 12 females). For receptive agrammatism, we used for comparison a group of 10 neurologically intact controls who completed the first sentence-to-picture matching task (mean age ± SD = 61.79 ± 7.69 years, range = 49 – 75 years; 6 females) and another group of 26 neurologically intact controls who completed the second sentence-to-picture matching task (mean age ± SD = 69.69 ± 5.77 years, range = 53 – 79 years; 18 females), both from our comprehensive speech-language assessment battery. For the syntax comprehension score and individual neuropsychological measurements, we resorted to previously-published normative data to calculate age-and education-adjusted Z-scores.^45^ Consistent with previous studies, Z-scores lower than −1.5 were considered abnormal.^45^ The output of these analyses allowed us to obtain a conservative estimate of the frequency of occurrence of expressive and receptive agrammatism across patients. For each clinical domain, a patient was labeled as having impaired performance if one or more of the scores assigned to that domain were abnormal (e.g., we considered that a patient had evidence of expressive agrammatism if either %UtWErrors, MLU, or SI were impaired). We used Venn diagrams to illustrate the frequency and partial overlap of distinct deficits.

### Classification of participants into clinically defined subgroups based on major speech and language characteristics

Participants with AOS but without expressive agrammatism were classified as PPAOS, while participants with expressive agrammatism but without AOS were classified as PAA according to previous work.^5, 6^ Participants with both AOS and expressive or receptive agrammatism meeting current consensus criteria^2^ were classified as AOS+agrammatism. Seven participants characterized by reduced speech fluency and dysarthria in the absence of AOS and agrammatism remained unclassifiable and were therefore assigned to a fourth group named "non/fluent dysarthric" (nf-Dysarthric).

### Assessing clustering tendency of raw data

We studied clustering tendency of raw (non-binary) data with the Hopkins statistic,^37^ to determine if participants with the nfvPPA-S tended to cluster into distinct clinically meaningful subgroups (i.e., a non-random structure). The Hopkins statistic represents the probability that a given dataset is generated by a uniform or random data distribution^46^ (values close to 0.5 indicate a low clustering tendency), while values far above 0.5 and close to 1 suggest the existence of clusters. We built hierarchical heatmaps to represent the patterns of individual performance of all participants. Hierarchical heatmaps have proven useful in identifying clusters of participants and clinical features within heterogeneous diseases.^47^ They consist of a colored matrix including individual data and clinical features, with the participants and the clinical features ordered by hierarchical clustering.

### Principal Component Analysis

Next, we adopted a data-driven approach to extract new LCDs. For this, we used Principal Component Analysis (PCA) in participants without missing values to avoid data imputation as a source of bias. Importantly, we employed PCA despite the observed low clustering tendency of the dataset because the existence (or lack) of clinically meaningful symptom clusters in a dataset does not equate to the existence (or lack) of underlying clinical dimensions with etiologic/prognostic value and vice versa. Based on Bartlett’s test of sphericity, the Kaiser–Meyer–Olkin value, and *a-priori* knowledge from the researchers, the following relevant features were entered into the PCA: apraxia of speech, dysarthria, speech fluency (words per minute), expressive agrammatism index, receptive agrammatism index, the non-verbal and verbal executive function indexes, and CDR-SB. By selecting these variables, the Bartlett’s test of sphericity (*χ*^²^(10) = 50.7, p < .001) and the Kaiser–Meyer–Olkin value (.71) indicated that the sample size and the correlation between the selected variables were suitable for PCA analysis.^48, 49^ Conversely, when including a higher number of features, the Bartlett’s test of sphericity and the Kaiser–Meyer–Olkin discouraged PCA analyses. Three components were selected based on Cattell’s criteria.^50^ To explore the potential value of data-driven LCDs to differentiate underlying etiology in our patients sample, we explored if the individual loadings of the extracted components (i.e., LCDs) allowed the discrimination between neuropathological subtypes in the subgroup of participants with autopsy data.

### Longitudinal analyses

Next, we investigated longitudinal decline with linear mixed-effects models. In longitudinal studies, linear mixed-effects models have proven to be powerful tools for identifying variables where baseline values are associated with different rates of change in clinical deterioration.^51^ We fitted a linear mixed-effects model controlling for age, sex, and the most relevant clinical features to estimate clinical deterioration over time (as measured by CDR-SB or CDR plus NACC FTLD-SB) across our patient group. We also fitted a linear mixed-effects model controlling for age, sex, and individual loadings for LCDs derived from the PCA analysis. All linear mixed-effects models included a random patient-specific intercept and a random patient-specific slope. These random effects account for patient heterogeneity in baseline CDR-SB (or CDR plus NACC FTLD-SB) and its rate of increase that is not explained by the predictors in the model. A term for clinical feature by time interaction was used to study the association between the baseline clinical feature and CDR-SB (or CDR plus NACC FTLD-SB) over time. As in similar previous studies,^52^ all linear mixed models were designed with a compound symmetry covariance structure (owing to the relative homogeneity in the covariance of effects). Of note, we obtained the same results when linear mixed models were fitted with unstructured covariance.

### Statistical analysis

We compared the raw clinical measurements between clinical subgroups in the whole sample using t-test or Kruskal–Wallis test for continuous variables and the Chi-square test for categorical data. We also compared the main clinical features between the groups of participants with and without autopsy data to verify that participants without autopsy data were clinically similar to those with a definitive neuropathological diagnosis. Next, in the subgroup of participants with an autopsy-proven diagnosis, we compared the clinical features between neuropathological categories with at least three participants (namely, PSP, CBD, Pick’s disease, and FTLD-TDP type B). To support the definition of speech, language, and cognitive domains, we explored the relationship between speech, language, and cognitive measures with Spearman’s correlation (for further details, please refer to the ’**Definition of speech, language, and cognitive domains’** section). Statistical significance for all tests was set at 5% (α = .05), all statistical tests were two-sided, and all group comparisons were corrected for multiple comparisons using the false discovery rate. All statistical analyses were conducted in R v4.1.1 (packages: ggplot2, ggstatsplot, ggVennDiagram, EFAtools, FactoMineR, lme4, ggeffects).

### Standard protocol approval, registration, and patient consent

The study was approved by the Institutional Review Board of UCSF and was conducted following the Declaration of Helsinki. All participants gave their written informed consent to participate in the study.

### Data availability

The conditions of our ethics approval do not permit the public archiving of anonymized study data. Data requests can be submitted here: https://memory.ucsf.edu/research-trials/professional/open-science. Following a UCSF-regulated procedure, access will be granted to designated individuals in line with ethical guidelines on reusing sensitive data. This would require the submission of a Material Transfer Agreement. Commercial use will not be approved.

## Results

### Baseline characteristics of the sample

**Table 1** shows the demographics and main clinical, genetic, and neuropathological features of the 98 participants included in this study. The mean (SD) age at diagnosis was 68.0 (7.1), and 63 participants (64%) were female. The mean (SD) time from estimated symptom onset to diagnosis was 4.3 (2.0) years, and almost 50% of the participants had a global score on the CDR plus NACC FTLD of 0.5 (indicating a mild/prodromal stage). A definitive neuropathological diagnosis was available in forty-three participants (44% of the sample). CBD and PSP were the two most frequently observed neuropathological diagnoses (n=17 [40%], and n=11 [26%], respectively). However, fifteen (34%) had other neuropathological diagnoses (**Table 1**). A pathogenic mutation in the *GRN* gene was found in 5 (5%) of the participants (3 of whom had autopsy confirmation of FTLD-TDP type A). Of note, participants with autopsy did not differ in terms of age at symptom onset, age at MRI, sex distribution, years of education, handedness distribution, disease severity, and NPI total score from participants without an autopsy-proven diagnosis (**Supplementary Tables 2-3**)

**Table 1.**
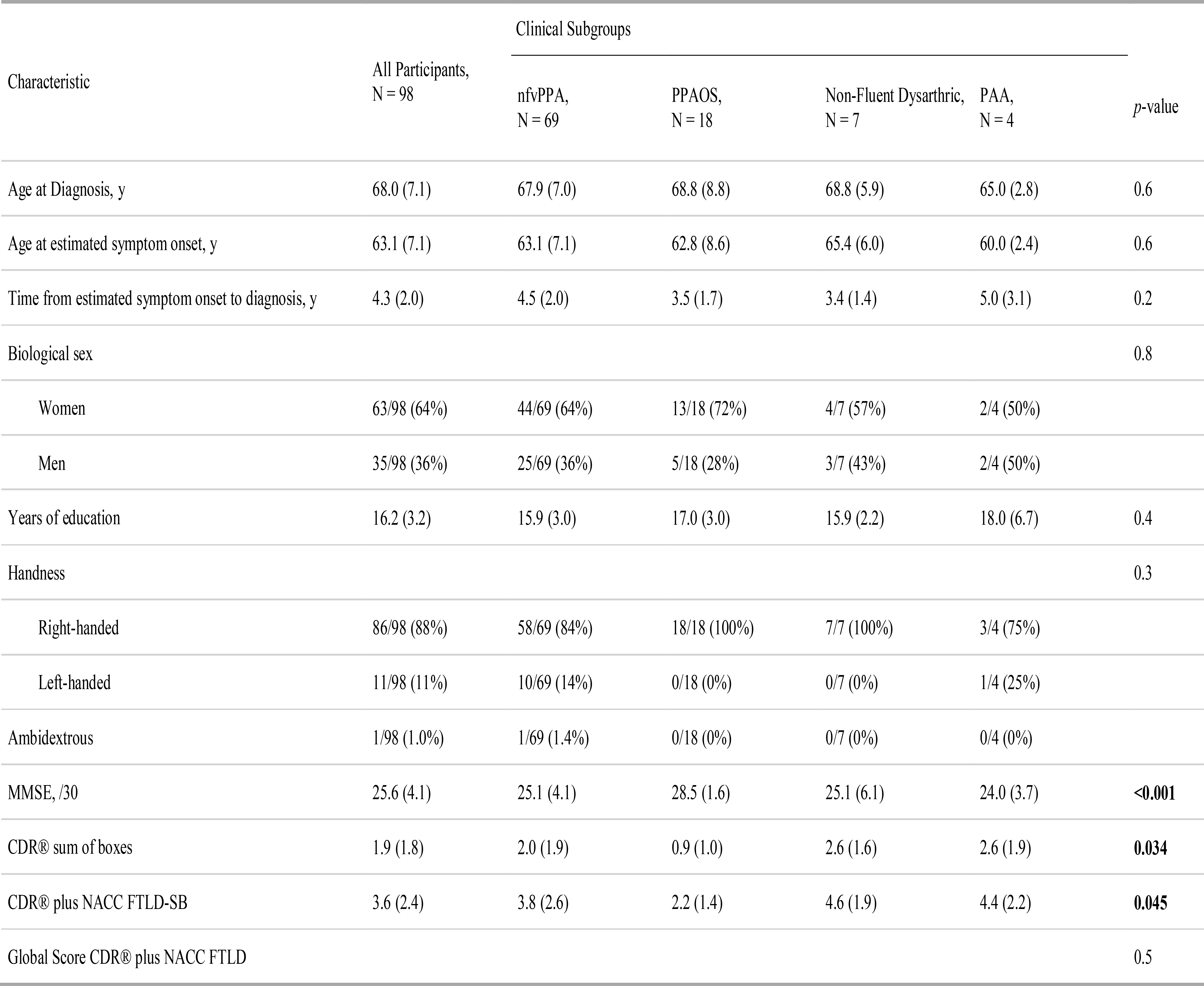
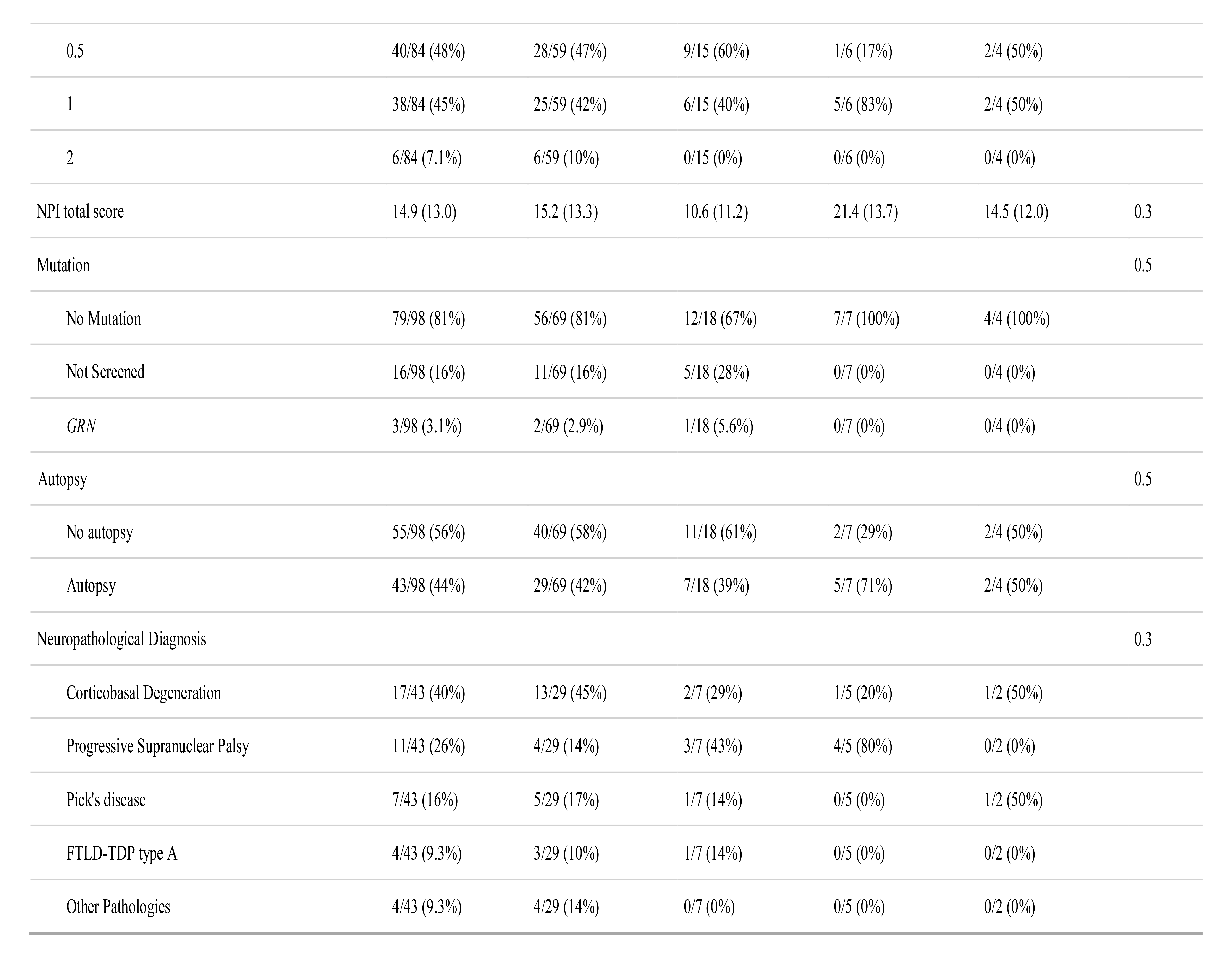
Demographic and clinical characteristics of participants with progressive speech and language disorders. Unless otherwise indicated, results are expressed as mean (SD). Statistically significant *p*-values are **bold**. The "non-fluent dysarthric" subgroup included seven participants with a variable combination of dysarthria and reduced speech fluency but without apraxia of speech or expressive agrammatism, not meeting previously reported criteria for any primary progressive speech-language syndrome. "Other Pathologies" included 1 participant with an unclassifiable tauopathy, 1 participant with AD, 1 with mixed AD and CBD, and 1 with FTLD-TDP type B. **Abbreviations:** CDR = clinical dementia rating; CDR® plus NACC FTLD-SB = clinical dementia rating plus national Alzheimer’s coordinating center frontotemporal lobar degeneration sum of boxes; FTLD = frontotemporal lobar degeneration; MMSE = mini-mental state examination, nfvPPA = non-fluent/agrammatic variant primary progressive aphasia; PPAOS = primary progressive apraxia of speech; PAA = primary agrammatic aphasia; NPI = neuropsychiatric inventory.

### Frequency of the main clinical features

As expected, the core features of nfvPPA were observed in a high proportion of participants (**Supplementary Table 4**). AOS and expressive agrammatism were present in 89% and 58% of the participants, respectively. Reduced speech fluency (words per minute) and reduced phonemic (letter) fluency were also frequently observed (91% and 84%, respectively). Overall, **Fig. 1** illustrates the significant overlap between the main speech, language, and cognitive features of participants within the nfvPPA-S. Motor speech deficits (either AOS or dysarthria) were noted in 97% of the participants (**Fig. 1A**), while agrammatism (either expressive or receptive) was observed in 75% (**Fig. 1B**). Notably, deficits in executive function were also detected in 66% of the participants when using non-verbal tests (**Supplementary Table 4**).

**Figure 1.**
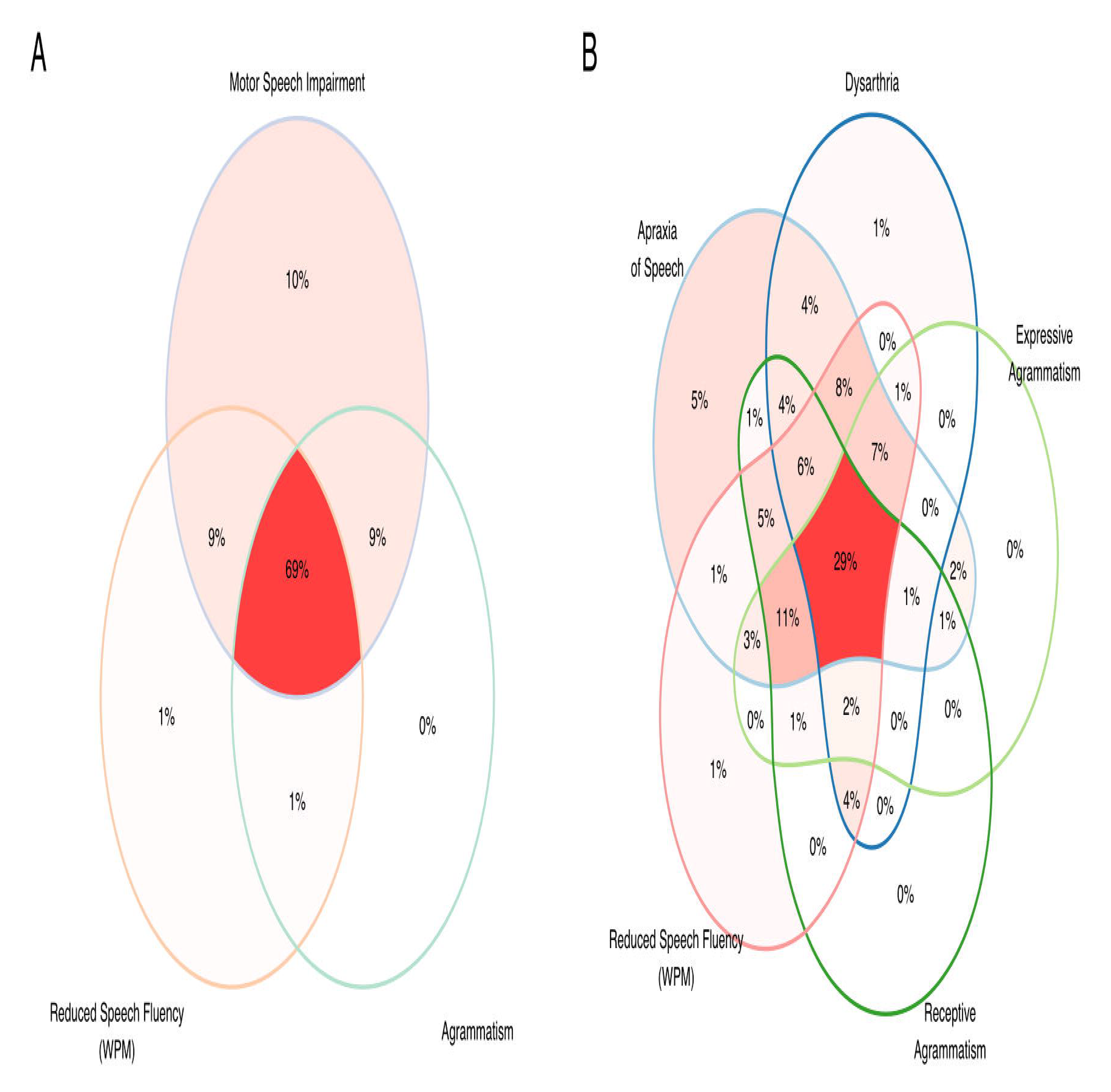
Frequency of main clinical features. Venn diagram illustrating the overlap between motor speech impairment, agrammatism, and reduced speech fluency **(A)**, apraxia of speech, dysarthria, expressive and receptive agrammatism, and reduced speech fluency (words per minute) **(B)**. **Abbreviations:** WPT=words per minute

### Frequency of previously reported clinical phenotypes

Across the nfvPPA-S, 69 participants (70%) had both AOS and agrammatism (either expressive or receptive), eighteen participants (18%) had apraxia of speech without agrammatism (PPAOS), while 4 participants (4%) had expressive agrammatism without AOS (PAA) (**Table 1**). Seven participants (7%) remained unclassifiable and were characterized by reduced speech fluency and dysarthria without AOS and agrammatism. These participants were assigned to a fourth group named "non/fluent dysarthric" (nf-Dysarthric).

### Comparison of clinical and neuropathological features between phenotypes

As shown in **Table 1**, the AOS+agrammatism, PPAOS, PPA, and nf-Dysarthric subgroups had similar ages at diagnosis and estimated time from symptom onset to diagnosis. However, participants in the PPAOS group were diagnosed at an earlier disease stage (as measured by the CDR-SB and the CDR® plus NACC FTLD-SB) and had better general cognitive performance (**Table 1** and **Supplementary Table 5**). Crucially, the AOS+agrammatism, PPAOS, PAA, and nf-Dysarthric subgroups were comparable regarding their neuropathological correlates (**Table 1**). Similarly, none of the baseline clinical features discriminated between FTLD subgroups after accounting for multiple comparisons (**Supplementary Table 6**).

### Cluster tendency analysis

Next, we explored the existence of clinically meaningful symptom clusters across our patient group. The clustered heatmap failed to reveal non-overlapping clusters of participants based on their major clinical features (**Fig. 2**). The lack of clustering tendency was confirmed by the Hopkins statistic (.454, p<.001). We also confirmed that neuropathological subtypes were similarly distributed along the nfvPPA-S rather than associated with specific clinical phenotypes, as illustrated in **Fig. 2**.

**Figure 2.**
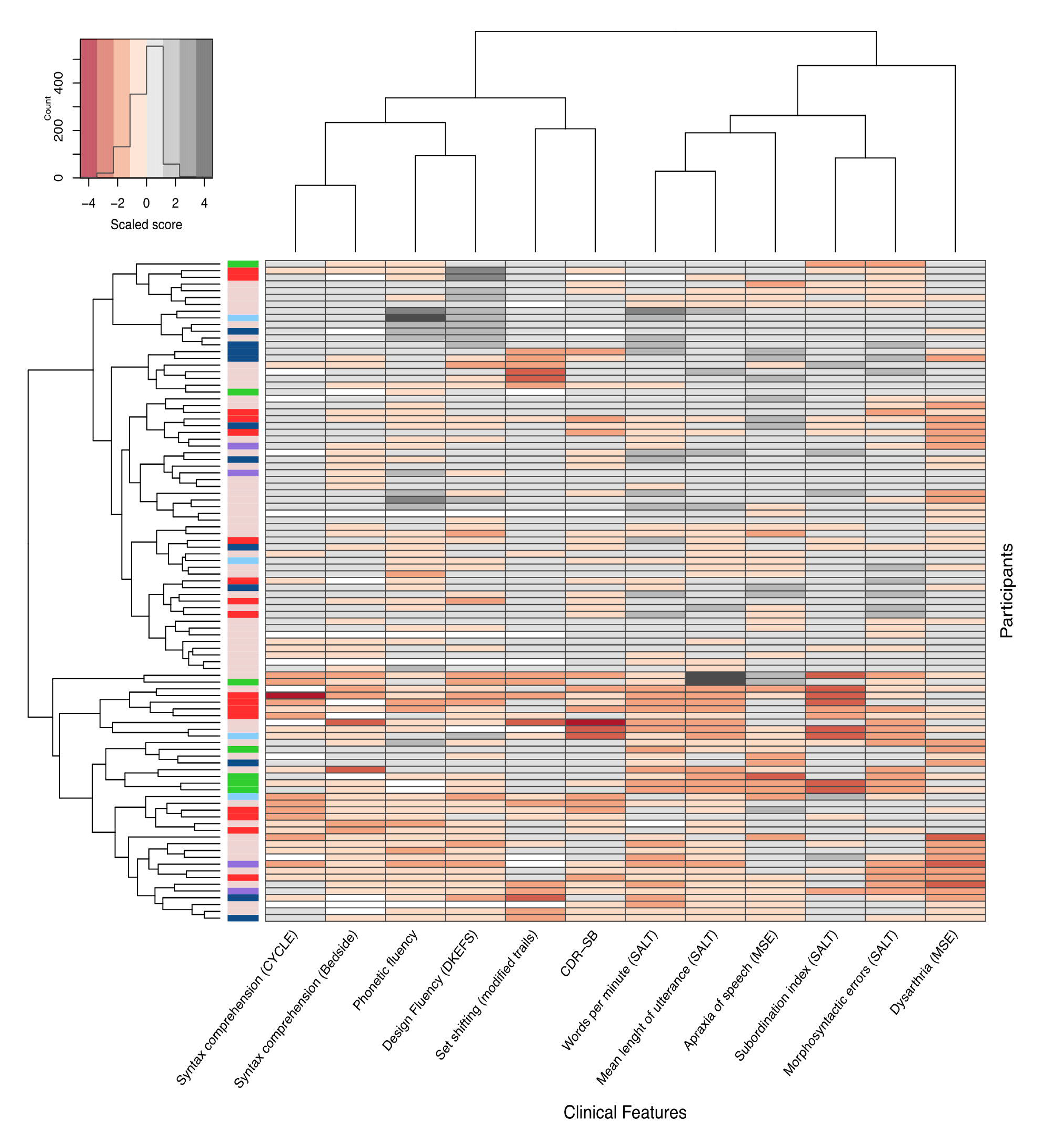
Clustered heatmap of all participants with progressive non-fluent speech-language disorders included in this study. The clustered heatmap illustrates the lack of robust clinical clusters within the nfvPPA-S. The first unlabeled column relates to the neuropathological data of each participant (pink = no autopsy available; red = CBD; dark blue = PSP; green = Pick’s disease; light blue = FTLD-TDP type A; purple = other pathologies). Each labeled column represents its corresponding clinical feature, and each row represents a participant. The scores for all clinical features have been scaled to allow their comparison. The participants and the variables have been ordered based on similarity, as illustrated by dendrograms on the top (variables) and left (participants). **Abbreviations:** SALT= Systematic Analysis of Language Transcripts; CYCLE= Curtiss Yamada Comprehensive Language Evaluation; MSE=Motor Speech Examination; CDR-SB= Clinical Dementia Rating sum-of-boxes; PSP=Progressive Supranuclear Palsy; CBD=Corticobasal Degeneration; FTLD-TDP= Frontotemporal Lobar Degeneration characterized by phosphorylated 43-kDa TAR DNA-binding protein inclusions; DKEFS= Delis-Kaplan Executive Function Scale.

### Principal component analysis

Because classifying participants into discrete clinical entities (i.e., syndromes) based on major speech and language characteristics was not useful for identifying the underlying pathology in our sample, we explored if PCA could unveil LCDs with etiologic value. Three LCDs explained 72% of the variance in the dataset. As shown in **Fig. 3**, the first component explained 37% of the variance and reflected reduced speech fluency (words per minute), executive dysfunction (both verbal and non-verbal), agrammatism (both expressive and receptive), and overall cognitive and functional impairment (as measured by the CDR-SB). The second component explained 19% of the variance and was defined by prominent AOS with dysarthria and lesser cognitive and functional impairment. The third component explained 12% of the variance and was mainly associated with dysarthria but not AOS.

**Figure 3.**
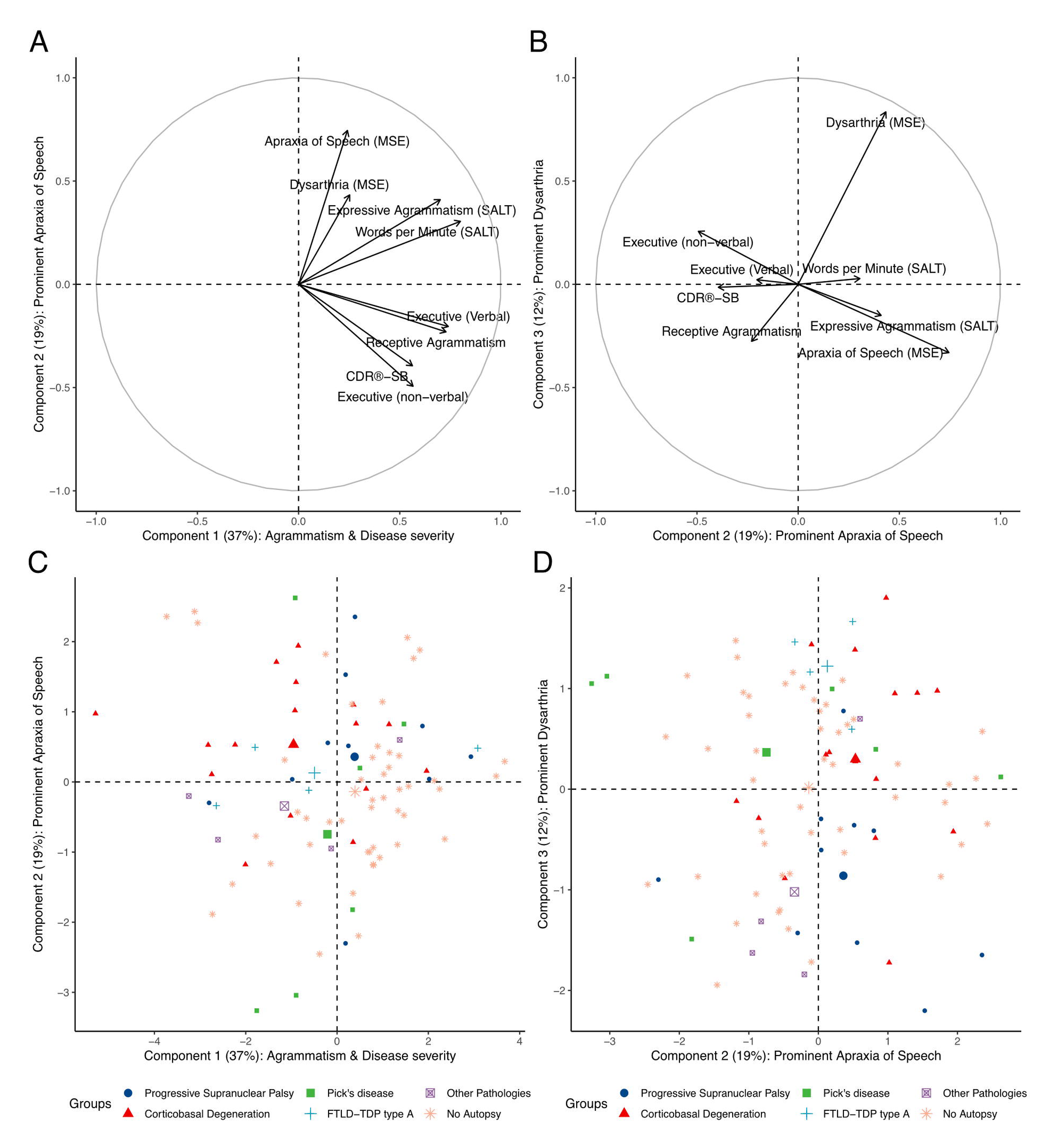
Characterization of latent clinical dimensions derived from principal component analysis. In all panels, the x-and y-axis represent one of the three latent clinical dimensions (or "clinical components") derived from PCA. The percentage of variance explained by each component is shown in parentheses. In panels **A)** and **B)**, the value at the x-and y-axis represent the standardized coefficient (relative weight) of the components derived from PCA (higher values indicating a more substantial contribution to a given component). Each variable included in the PCA is represented by an arrow whose orthogonal projection to the x-and the y-axis indicates its standardized coefficient for the corresponding component. Panel **A)** illustrates the relative contribution of each variable to Component 1 (characterized by agrammatism, reduced verbal fluency, and higher disease severity) and Component 2 (represented by prominent apraxia of speech and less disease severity and executive dysfunction). Panel **B)** illustrates the relative contribution of each variable to Component 2 and Component 3 (characterized by prominent dysarthria with less apraxia of speech). Panels **C)** and **D)** represent the individual loadings for each component (lower individual loadings representing more impaired performance). Each participant’s neuropathological diagnosis (if available) is shown in the legend of panels **C)** and **D).** For each neuropathological category, the centroid of the ellipse encompassing each group is represented by the biggest symbol. Notably, the distribution of individual loadings did not reveal clusters of participants but rather a widespread distribution. In panels **C)** and **D)**, the x-and y-axis represent the individual loading for one of the three components derived from PCA Panels **C)** and **D**). **Abbreviations:** LCD=latent clincal dimension; PCA= Principal Component Analysis; CDR-SB= Clinical Dementia Rating sum-of-boxes;

Next, we explored the neuropathological correlates of the three LCDs (from the PCA) in the subgroup of participants with autopsy data. As shown in **Fig. 4**, the first and second dimensions did not yield significant differences between the neuropathological subgroups (**4A and 4B**). However, the LCD reflecting prominent dysarthria was significantly reduced in participants with PSP when compared to participants with CBD, Pick’s disease, or FTLD-TDP type A.

**Figure 4.**
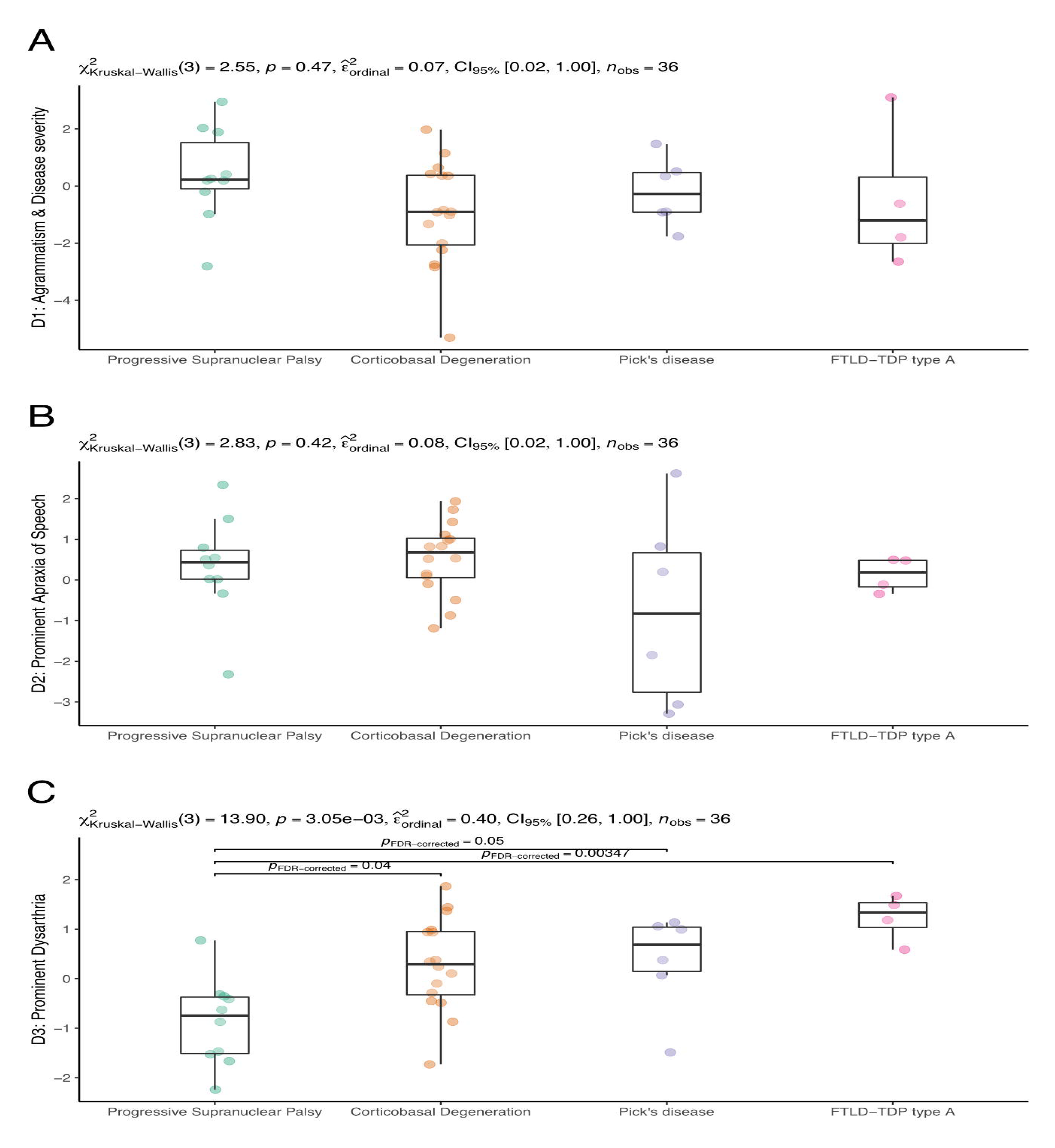
Neuropathological correlates of data-driven latent clinical dimensions. Subject-specific loading for each latent clinical component was available in 85 (83%) participants from the total sample and 36 (84%) participants with neuropathological diagnoses. **Abbreviations:** D1=latent clinical dimension 1; D2= latent clinical dimension 2; D3= latent clinical dimension 3

### Longitudinal analyses

When considering baseline raw data, higher scores for dysarthria, reduced speech fluency (words per minute), and higher expressive and receptive agrammatism were associated with an increased rate of clinical decline as measured with both the CDR-SB and the CDR plus NACC FTLD-SB (**Supplementary Table 7**). When considering the components derived from the PCA (**Supplementary Table 8 and Fig. 5A**), only the LCD characterized by higher disease severity and worst agrammatism was associated with an increased rate of clinical decline (**Fig. 5B**). Of note, the LCD characterized by prominent AOS was associated with lower disease severity at baseline, supporting the view that participants with relatively isolated AOS are typically diagnosed at an earlier disease stage than the rest of the participants.

**Figure 5.**
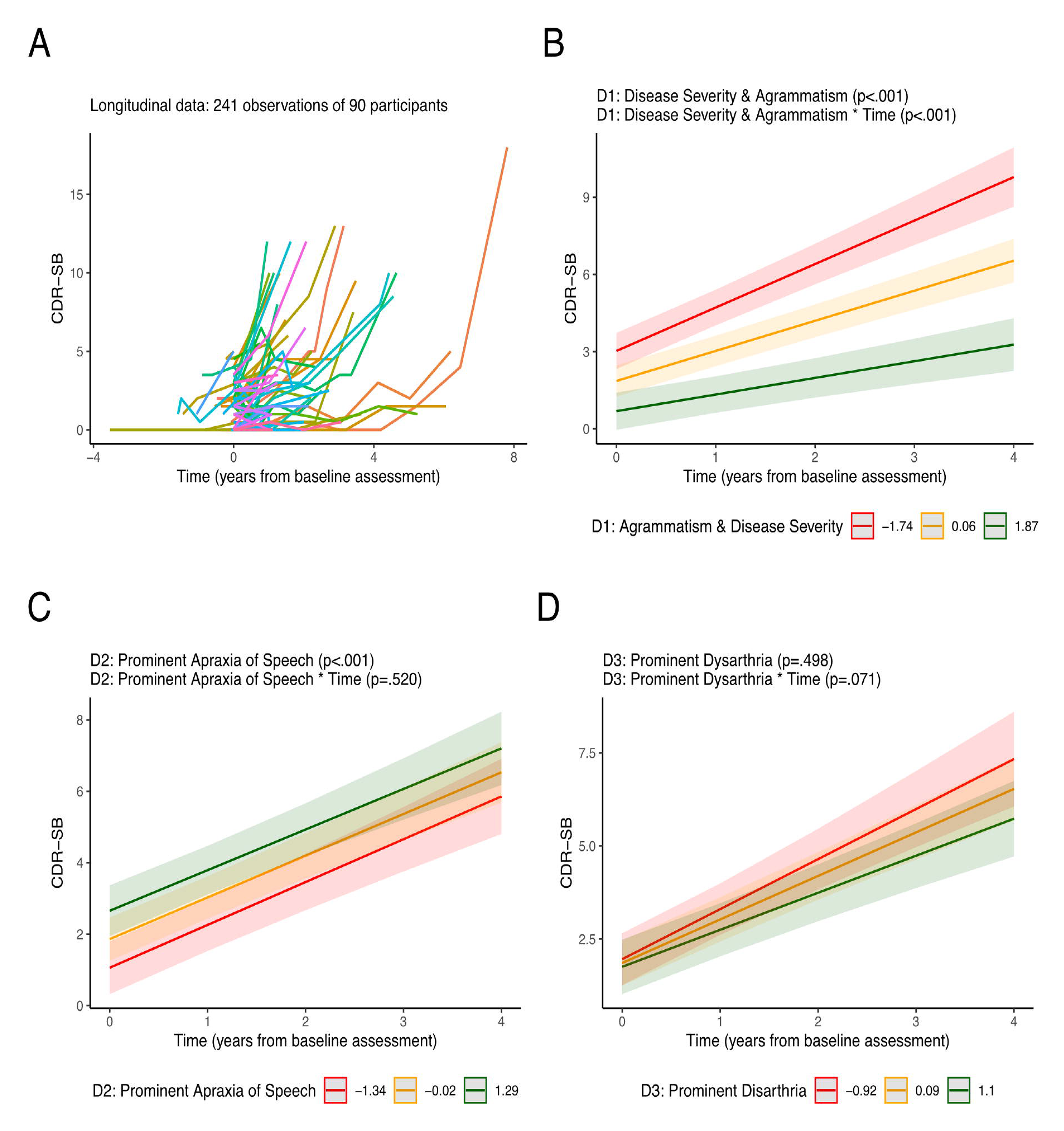
Baseline predictors of faster longitudinal decline. **A)** Spaghetti plot representing longitudinal data. **B-D)** CDR-SB estimates were obtained from linear mixed-effects models as a function of individual loading for Components 1, 2, and 3. For illustrative purposes, we show the CDR-SB estimates for each tercile. Error bars represent 95% confidence intervals, and lower values for each latent clinical component represent more impaired performance. **Abbreviations:** D1=component 1; D2=component 2; D3=component 3; CDR-SB= Clinical Dementia Rating sum-of-boxes

## Discussion

In this study, we leveraged a large cohort of participants within the non-fluent primary progressive aphasia spectrum to examine the presence of robust endophenotypes and their potential to enhance clinical-pathological correlations and prognosticate disease progression. Our findings indicate that this clinical entity embodies a continuous spectrum of substantially overlapping cognitive, speech, and language characteristics. Although data-driven exploration of LCDs did not prove beneficial in predicting the underlying pathology, the clinical dimension characterized by a reduced words-per-minute rate, agrammatism, and increased cognitive and functional impairments emerged as a significant predictor of a more rapid rate of decline.

A noteworthy aspect of the present study involves the objective quantification of expressive and receptive agrammatism in an extensive cohort of participants within the nfvPPA-S. The multifaceted nature of grammatical processing renders it challenging to assess in routine clinical practice. The absence of standardized tests for evaluating expressive and receptive agrammatism and discrepancies in defining the threshold for "significant" agrammatism may account for the variation in reported prevalence observed in prior studies. In our sample, up to 75% of participants demonstrated varying degrees of either expressive or receptive agrammatism. Concurrently, motor speech deficits, such as apraxia of speech (AOS) or dysarthria, were nearly universal. We identified two distinct subgroups among the subset of participants who presented exclusively with motor speech impairments (25%) at baseline. The first subgroup exhibited marked AOS, meeting the diagnostic criteria for primary progressive apraxia of speech (PPAOS). In contrast, the second subgroup was characterized by pronounced dysarthria and additional clinical features, including reduced speech fluency and executive dysfunction. This latter group remained unclassifiable per current diagnostic criteria and was predominantly observed in patients with progressive supranuclear palsy (PSP), with 4/5 (80%) of participants in this group having autopsy-confirmed PSP. We recognize that subtle differences in the subjective evaluation of apraxia of speech and dysarthria (**Supplementary Table 1**) may influence participant classification. Further studies are therefore needed to investigate the degree to which the classification of AOS (including potential subtypes) and dysarthria, based on expert-driven auditory-perceptual assessments, can be replicated across various research centers and countries.

Another critical finding of our study is that participants with nfvPPA-S presentation cannot be robustly clustered into separate clinical syndromes (e.g., AOS+agrammatism, PPAOS, PAA) but rather fall along a clinical spectrum with substantial overlap behaviorally and pathologically. Along this clinical continuum, we found that the LCD characterized by disease severity and agrammatism was an important prognostic factor. Participants with expressive agrammatism and a more severe cognitive and functional impairment (as measured by CDR-SB) were characterized by a faster rate of clinical decline. This finding supports the view that reduced speech fluency, executive dysfunction, and general functional impairment should be considered in tandem with agrammatism to stratify participants based on their progression rate. Such stratification may reduce the heterogeneity of participants in future trials and reduce sample size requirements.^53^

The finding that a faster disease progression characterizes participants presenting with marked agrammatism is also relevant for interpreting previous studies. For example, in keeping with our results, it has been reported that patients with prominent expressive agrammatism in the absence of AOS (referred to as "primary agrammatic aphasia", PAA) display faster disease progression and more widespread neurodegeneration during follow-up.^5^ Conversely, participants in our sample classified as PPAOS (AOS in the absence of expressive agrammatism) showed milder disease severity, as measured with the CDR-SB (and the CDR® plus NACC FTLD-SB) than the rest of the participants. This finding suggests that participants with relatively isolated AOS may be diagnosed at an earlier disease stage than those with relatively isolated agrammatism. This is an important observation because disease progression has been shown to accelerate with increasing disease severity in FTLD.^54^ Thus, patients diagnosed at a more advanced disease stage are expected to deteriorate faster than those diagnosed at an earlier disease stage. Accordingly, our results support the view that patients with significant agrammatism at diagnosis may be at a more advanced disease stage and are thus expected to deteriorate faster. Of note, validated staging tools, such as the CDR® plus NACC FTLD-SB, are more likely to yield reproducible results than the widely adopted “estimated time from symptom onset”, which is a retrospective measure influenced by recall bias, patient anosognosia and other patient-and clinician-related factors.^32^

Consistent with the lack of robust clustering tendency in our large and representative dataset, we showed that classifying participants into supposedly clear-cut clinical phenotypes (i.e., PPAOS vs PAA) based on their core clinical features did not predict underlying pathology. This finding prompted the investigation of data-driven components to improve clinical-pathological correlations and prediction of disease progression. Our data-driven approach in the nfvPPA-S contrasts with subjective expert-based perceptual classifications and provides valuable information to interpret previous studies.^9^ We found that the LCD characterized by prominent dysarthria distinguished participants with underlying PSP from those with other neuropathological subtypes. This finding is in line with previous studies from our group.^14^ However, we also observed substantial overlap between clinical features, which, alone, did not allow the prediction of underlying etiology at the single-subject level. It should be noted, however, that segregating participants according to their clinical profile for treatment purposes may still be useful, as different patients may benefit from some speech therapy interventions but not others depending on their characteristic deficits.^55^ However, as previously mentioned, most current approaches include strategies for both motor speech and grammatical deficits that are likely to occur with disease progression.

As expected, the nfvPPA-S was associated with multiple neuropathological diagnoses. In keeping with previous studies, 4R tauopathies (particularly PSP) were frequently observed in participants with AOS but not agrammatism. However, the data-driven LCD defined in this study did not allow robust discrimination of neuropathological subtypes. It should be noted that the diagnosis of AOS and dysarthria (two motor speech disorders) is based on expert-dependent auditory-perceptual assessments that are prone to systematic bias and noise. Thus, robust and reproducible quantitative, objective measures of AOS and other motor speech impairments are needed to advance the classification of the nfvPPA-S. For example, in a recent study from our group, we showed that automated speech timing measures (i.e., articulation rate) might provide valuable information to characterize the nfvPPA-S and its neuropathological bases.^56^ Thanks to its objectivity and scalability, automated speech analysis could offer important diagnostic support to standard speech assessments. However, further studies are needed to determine these new measurements’ full diagnostic and prognostic potential.

Taken together, our result do not support the view that the nfvPPA-S can be robustely split into clear-cut syndromic entities (i.e., PPAOS vs PAA) or that these supposedly clear-cut clinical phenotypes allow discriminating between FTLD pathological subtypes. A current challenge to the field is that FTLD-related syndromes are largely overlapping, and many patients may meet diagnostic criteria for more than one syndrome.^3^ For example, the distribution of different neuropathological entities in our cohort is very similar to that reported in a cohort of PPAOS patients (**Supplementary Fig.**), suggesting that these two diagnostic labels are largely overlapping.^9^ More work is thus needed to implement multidimensional classification schemes that incorporate imaging^57, 58^ and fluid biomarkers to advance toward precision medicine approaches to pharmacological treatment. Whether focusing early cognitive therapy on the early predominant symptom or supporting functions likely to decline later on remains to be established.

This study has some limitations. For example, a definitive neuropathological diagnosis was only available in a relatively small number of participants (n = 43), limiting statistical power. Yet, to the best of our knowledge, our study still represents the largest series of cases with autopsy data, amounting to nearly half of the total sample. In addition, the measure we derived for expressive agrammatism (from a quantitative linguistic analysis of connected speech samples) may miss some cases with milder/subtler forms of expressive agrammatism that trained experts could detect. Finally, our dataset only included semiquantitative measures of AOS and dysarthria obtained from an expert-dependent auditory-perceptual assessment of motor speech ability. Future studies should also consider additional quantitative measures derived from expert-independent automated speech analyses.

Splitting the nfvPPA-S disorder into different clinical phenotypes based on its major clinical features does not improve clinical-pathological correlations, stressing the need for new clinical and biological markers.

## Acknowledgments

We thank the patients and their families for participation in this research.

## Supplementary Material

### Supplementary Tables

**Supplementary Table 1.** Motor speech characteristics indicative of apraxia of speech or dysarthria considered in this study.

**Supplementary Table 2.** Demographic and clinical characteristics of the participants with and without autopsy.

**Supplementary Table 3.** Speech, language, and cognitive features of the participants with and without autopsy.

**Supplementary Table 4.** Frequency of main speech, language, and cognitive features.

**Supplementary Table 5.** Comparison of clinical features between clinical subgroups.

**Supplementary Table 6.** Comparison of clinical features between neuropathological groups.

**Supplementary Table 7.** Linear Mixed Effects Model for the estimation of CDR-SB based on raw clinical data.

**Supplementary Table 8.** Linear Mixed Effects Model for the estimation of CDR-SB based on latent clinical dimensions.

### Supplementary Figures

**Supplementary Fig. 1.** Neuropathological diagnoses along the spectrum of progressive non-fluent speech and language disorders.

## References

1. Tee BL, Gorno-Tempini ML. Primary progressive aphasia: a model for neurodegenerative disease. Curr Opin Neurol. 2019;32(2):255–265.

2. Gorno-Tempini ML, Hillis AE, Weintraub S, et al. Classification of primary progressive aphasia and its variants. Neurology. 2011;76(11):1006–1014. doi:10.1212/WNL.0b013e31821103e6

3. Murley AG, Coyle-Gilchrist I, Rouse MA, et al. Redefining the multidimensional clinical phenotypes of frontotemporal lobar degeneration syndromes. Brain. 2020;143(5):1555–1571. doi:10.1093/brain/awaa097

4. Josephs KA, Duffy JR, Strand EA, et al. The evolution of primary progressive apraxia of speech. Brain. 2014;137(10):2783–2795.

5. Tetzloff KA, Duffy JR, Clark HM, et al. Progressive agrammatic aphasia without apraxia of speech as a distinct syndrome. Brain. 2019;142(8):2466–2482. doi:10.1093/brain/awz157

6. Josephs KA, Duffy JR, Strand EA, et al. Characterizing a neurodegenerative syndrome: primary progressive apraxia of speech. Brain. 2012;135(5):1522–1536.

7. Boeve BF, Boxer AL, Kumfor F, Pijnenburg Y, Rohrer JD. Advances and controversies in frontotemporal dementia: diagnosis, biomarkers, and therapeutic considerations. Lancet Neurol. 2022;21(3):258–272. doi:10.1016/S1474-4422(21)00341-0

8. Thompson CK, Cho S, Price C, et al. Semantic interference during object naming in agrammatic and logopenic primary progressive aphasia (PPA). Brain Lang. 2012;120(3):237–250. doi:10.1016/j.bandl.2011.11.003

9. Josephs KA, Duffy JR, Clark HM, et al. A molecular pathology, neurobiology, biochemical, genetic and neuroimaging study of progressive apraxia of speech. Nat Commun. 2021;12(1):3452. doi:10.1038/s41467-021-23687-8

10. Henry ML, Hubbard HI, Grasso SM, et al. Retraining speech production and fluency in non-fluent/agrammatic primary progressive aphasia. Brain J Neurol. 2018;141(6):1799–1814. doi:10.1093/brain/awy101

11. Spinelli EG, Mandelli ML, Miller ZA, et al. Typical and atypical pathology in primary progressive aphasia variants. Ann Neurol. 2017;81(3):430–443.

12. Whitwell JL, Tosakulwong N, Schwarz CC, et al. Longitudinal anatomic, functional, and molecular characterization of Pick disease phenotypes. Neurology. 2020;95(24):e3190–e3202. doi:10.1212/WNL.0000000000010948

13. Caso F, Mandelli ML, Henry M, et al. In vivo signatures of nonfluent/agrammatic primary progressive aphasia caused by FTLD pathology. Neurology. 2014;82(3):239–247.

14. Santos-Santos MA, Mandelli ML, Binney RJ, et al. Features of Patients With Nonfluent/Agrammatic Primary Progressive Aphasia With Underlying Progressive Supranuclear Palsy Pathology or Corticobasal Degeneration. JAMA Neurol. 2016;73(6):733–10.

15. Rogalski E, Sridhar J, Rader B, et al. Aphasic variant of Alzheimer disease: Clinical, anatomic, and genetic features. Neurology. 2016;87(13):1337–1343.

16. Josephs KA. Clinicopathological and imaging correlates of progressive aphasia and apraxia of speech. Brain. 2006;129(6):1385–1398. doi:10.1093/brain/awl078

17. Boxer AL, Gold M, Feldman H, et al. New directions in clinical trials for frontotemporal lobar degeneration: Methods and outcome measures. Alzheimers Dement. 2020;16(1):131–143. doi:10.1016/j.jalz.2019.06.4956

18. Strafella C, Caputo V, Galota MR, et al. Application of Precision Medicine in Neurodegenerative Diseases. Front Neurol. 2018;9:701. doi:10.3389/fneur.2018.00701

19. Matias-Guiu JA, Díaz-Álvarez J, Cuetos F, et al. Machine learning in the clinical and language characterisation of primary progressive aphasia variants. Cortex J Devoted Study Nerv Syst Behav. 2019;119:312–323. doi:10.1016/j.cortex.2019.05.007

20. Hoffman P, Sajjadi SA, Patterson K, Nestor PJ. Data-driven classification of patients with primary progressive aphasia. Brain Lang. 2017;174:86–93. doi:10.1016/j.bandl.2017.08.001

21. Mesulam MM. Primary progressive aphasia. Ann Neurol. 2001;49(4):425–432.

22. Folstein MF, Folstein SE, McHugh PR. “Mini-mental state.” J Psychiatr Res. 1975;12(3):189–198. doi:10.1016/0022-3956(75)90026-6

23. Morris JC. The Clinical Dementia Rating (CDR): current version and scoring rules. Neurology. 1993;43(11):2412–2414. doi:10.1212/wnl.43.11.2412-a

24. Litvan I, Agid Y, Calne D, et al. Clinical research criteria for the diagnosis of progressive supranuclear palsy (Steele-Richardson-Olszewski syndrome): Report of the NINDS-SPSP International Workshop. Neurology. 1996;47(1):1–9. doi:10.1212/WNL.47.1.1

25. Boxer AL, Geschwind MD, Belfor N, et al. Patterns of Brain Atrophy That Differentiate Corticobasal Degeneration Syndrome From Progressive Supranuclear Palsy. Arch Neurol. 2006;63(1):81. doi:10.1001/archneur.63.1.81

26. Perry DC, Brown JA, Possin KL, et al. Clinicopathological correlations in behavioural variant frontotemporal dementia. Brain J Neurol. 2017;140(12):3329–3345. doi:10.1093/brain/awx254

27. Seo SW, Thibodeau MP, Perry DC, et al. Early vs late age at onset frontotemporal dementia and frontotemporal lobar degeneration. Neurology. 2018;90(12):e1047–e1056.

28. Mackenzie IRA, Neumann M, Bigio EH, et al. Nomenclature and nosology for neuropathologic subtypes of frontotemporal lobar degeneration: an update. Acta Neuropathol (Berl). 2010;119(1):1–4. doi:10.1007/s00401-009-0612-2

29. Gorno-Tempini ML, Brambati SM, Ginex V, et al. The logopenic/phonological variant of primary progressive aphasia. Neurology. 2008;71(16):1227–1234.

30. Wilson SM, DeMarco AT, Henry ML, et al. Variable disruption of a syntactic processing network in primary progressive aphasia. Brain. 2016;139(11):2994–3006. doi:10.1093/brain/aww218

31. Kramer JH, Jurik J, Sha SJ, et al. Distinctive neuropsychological patterns in frontotemporal dementia, semantic dementia, and Alzheimer disease. Cogn Behav Neurol Off J Soc Behav Cogn Neurol. 2003;16(4):211–218. doi:10.1097/00146965-200312000-00002

32. Miyagawa T, Brushaber D, Syrjanen J, et al. Utility of the global CDR® plus NACC FTLD rating and development of scoring rules: Data from the ARTFL/LEFFTDS Consortium. Alzheimers Dement J Alzheimers Assoc. 2020;16(1):106–117. doi:10.1002/alz.12033

33. Wertz RT, LaPointe LL, Rosenbek JC. Apraxia of Speech in Adults: The Disorder and Its Management. Grune & Stratton; 1984.

34. Kertesz A. The Western Aphasia Battery. Grune [and] Stratton; 1982.

35. Dronkers NF, Wilkins DP, Van Valin RD, Redfern BB, Jaeger JJ. Lesion analysis of the brain areas involved in language comprehension. Cognition. 2004;92(1-2):145–177. doi:10.1016/j.cognition.2003.11.002

36. Wilson SM, Dronkers NF, Ogar JM, et al. Neural Correlates of Syntactic Processing in the Nonfluent Variant of Primary Progressive Aphasia. J Neurosci. 2010;30(50):16845–16854. doi:10.1523/JNEUROSCI.2547-10.2010

37. Yi A. California Verbal Learning Test (California Verbal Learning Test-II). In: Kreutzer JS, DeLuca J, Caplan B, eds. Encyclopedia of Clinical Neuropsychology. Springer New York; 2011:475–476. doi:10.1007/978-0-387-79948-3_1112

38. Possin KL, Laluz VR, Alcantar OZ, Miller BL, Kramer JH. Distinct neuroanatomical substrates and cognitive mechanisms of figure copy performance in Alzheimer’s disease and behavioral variant frontotemporal dementia. Neuropsychologia. 2011;49(1):43–48. doi:10.1016/j.neuropsychologia.2010.10.026

39. Golden CJ, Freshwater SM, Zarabeth G, University NS. Stroop Color and Word Test Children’s Version for Ages 5-14: A Manual for Clinical and Experimental Uses. Stoelting; 2003. https://books.google.es/books?id=WyHwkQEACAAJ

40. Delis DC, Kaplan E, Kramer JH. Delis Kaplan Executive Function System (D-KEFS). Psychological Corporation; 2001. https://books.google.es/books?id=mnQqPwAACAAJ

41. Reitan RM. Validity of the Trail Making Test as an Indicator of Organic Brain Damage. Percept Mot Skills. 1958;8(3):271–276. doi:10.2466/pms.1958.8.3.271

42. Mack WJ, Freed DM, Williams BW, Henderson VW. Boston Naming Test: shortened versions for use in Alzheimer’s disease. J Gerontol. 1992;47(3):P154–158. doi:10.1093/geronj/47.3.p154

43. Cummings JL, Mega M, Gray K, Rosenberg-Thompson S, Carusi DA, Gornbein J. The Neuropsychiatric Inventory: Comprehensive assessment of psychopathology in dementia. Neurology. 1994;44(12):2308–2308. doi:10.1212/WNL.44.12.2308

44. Crawford JR, Garthwaite PH, Ryan K. Comparing a single case to a control sample: testing for neuropsychological deficits and dissociations in the presence of covariates. Cortex J Devoted Study Nerv Syst Behav. 2011;47(10):1166–1178. doi:10.1016/j.cortex.2011.02.017

45. Ranasinghe KG, Rankin KP, Lobach IV, et al. Cognition and neuropsychiatry in behavioral variant frontotemporal dementia by disease stage. Neurology. 2016;86(7):600–610. doi:10.1212/WNL.0000000000002373

46. Lawson RG, Jurs PC. New index for clustering tendency and its application to chemical problems. J Chem Inf Comput Sci. 1990;30(1):36–41. doi:10.1021/ci00065a010

47. Gómez-Andrés D, Oulhissane A, Quijano-Roy S. Two decades of advances in muscle imaging in children: from pattern recognition of muscle diseases to quantification and machine learning approaches. Neuromuscul Disord. 2021;31(10):1038–1050. doi:10.1016/j.nmd.2021.08.006

48. Bartlett MS. THE EFFECT OF STANDARDIZATION ON A χ 2 APPROXIMATION IN FACTOR ANALYSIS. Biometrika. 1951;38(3-4):337-344. doi:10.1093/biomet/38.3-4.337

49. Kaiser HF, Rice J. Little Jiffy, Mark Iv. Educ Psychol Meas. 1974;34(1):111-117. doi:10.1177/001316447403400115

50. Field AP, Miles J, Field Z. Discovering Statistics Using R. Sage; 2012.

51. Staffaroni AM, Ljubenkov PA, Kornak J, et al. Longitudinal multimodal imaging and clinical endpoints for frontotemporal dementia clinical trials. Brain J Neurol. 2019;142(2):443–459. doi:10.1093/brain/awy319

52. Illán-Gala I, Falgàs N, Friedberg A, et al. Diagnostic Utility of Measuring Cerebral Atrophy in the Behavioral Variant of Frontotemporal Dementia and Association With Clinical Deterioration. JAMA Netw Open. 2021;4(3):e211290. doi:10.1001/jamanetworkopen.2021.1290

53. Staffaroni AM, Weintraub S, Rascovsky K, et al. Uniform data set language measures for bvFTD and PPA diagnosis and monitoring. Alzheimers Dement Amst Neth. 2021;13(1):e12148. doi:10.1002/dad2.12148

54. Staffaroni AM, Goh SYM, Cobigo Y, et al. Rates of Brain Atrophy Across Disease Stages in Familial Frontotemporal Dementia Associated With MAPT, GRN, and C9orf72 Pathogenic Variants. JAMA Netw Open. 2020;3(10):e2022847. doi:10.1001/jamanetworkopen.2020.22847

55. Robinaugh G, Henry ML. Behavioral interventions for primary progressive aphasia. Handb Clin Neurol. 2022;185:221–240. doi:10.1016/B978-0-12-823384-9.00011-6

56. García AM, Welch AE, Mandelli ML, et al. Automated Detection of Speech Timing Alterations in Autopsy-Confirmed Non-fluent/agrammatic Variant Primary Progressive Aphasia. Neurology. Published online May 27, 2022:10.1212/WNL.0000000000200750. doi:10.1212/WNL.0000000000200750

57. Illán-Gala I, Nigro S, VandeVrede L, et al. Diagnostic Accuracy of Magnetic Resonance Imaging Measures of Brain Atrophy Across the Spectrum of Progressive Supranuclear Palsy and Corticobasal Degeneration. JAMA Netw Open. 2022;5(4):e229588. doi:10.1001/jamanetworkopen.2022.9588

58. VandeVrede L, Ljubenkov PA, Rojas JC, Welch AE, Boxer AL. Four-Repeat Tauopathies: Current Management and Future Treatments. Neurotherapeutics. 2020;17(4):1563–1581. doi:10.1007/s13311-020-00888-5

