## Supplementary Material for "Clinical dimensions along the progressive nonfluent variant primary progressive aphasia spectrum"

**Supplementary Tables**

**Supplementary Table 1. Motor speech characteristics indicative of apraxia of speech or dysarthria considered in this study**

| Apraxia of Speech | Spastic Dysarthria | Hypokinetic Dysarthria |
| --- | --- | --- |
| Slow speech rate  Distorted articulation  Distorted sound substitutions and/or additions  Sound sequencing errors  Articulatory groping/false starts  Trial-and-error articulation  Difficulty initiating speech  Reduced accuracy with increased utterance length, complexity and/or rate  Prosodic alterations | Slow speech rate  Strained-strangled/harsh voice quality  Imprecise articulation  Audible/strenuous  inspiration  Hypernasality  Slow, regular speech alternating motion rates  Pitch breaks | Accelerated speech rate  Breathy/harsh voice quality  Short rushes of speech  Imprecise articulation  Reduced loudness and stress  Inappropriate silences  Repeated sounds  Rapid, blurred speech alternating motion rates  Monopitch and  monoloudness |

**Supplementary Table 2. Demographic and clinical characteristics of the participants with and without autopsy.**

| **Characteristic** | **N** | **All Participants,**  **N = 98^1^** | **No autopsy,**  **N = 55^1^** | **Autopsy,**  **N = 43^1^** | **p-value** |
| --- | --- | --- | --- | --- | --- |
| **Age at diagnosis, y** | **98** | **68.2 (62.9, 73.8)** | **68.2 (63.0, 72.7)** | **68.8 (63.3, 74.9)** | **0.72** |
| **Age at estimated symptom onset, y** | **83** | **62.0 (58.5, 68.0)** | **62.0 (57.0, 66.0)** | **62.5 (59.2, 68.8)** | **0.42** |
| **Time from estimated symptom onset to diagnosis, y** | **83** | **4.1 (2.8, 5.2)** | **4.2 (2.9, 5.1)** | **3.9 (2.8, 5.4)** | **0.93** |
| **Biological sex** | **98** |  |  |  | **0.124** |
| **Women** |  | **63/98 (64%)** | **39/55 (71%)** | **24/43 (56%)** |  |
| **Men** |  | **35/98 (36%)** | **16/55 (29%)** | **19/43 (44%)** |  |
| **Years of education** | **98** | **16.0 (14.0, 18.0)** | **16.0 (14.0, 18.0)** | **16.0 (14.0, 18.0)** | **0.62** |
| **Handness** | **98** |  |  |  | **0.35** |
| **Right handed** |  | **86/98 (88%)** | **46/55 (84%)** | **40/43 (93%)** |  |
| **Left handed** |  | **11/98 (11%)** | **8/55 (15%)** | **3/43 (7.0%)** |  |
| **Ambidextrous** |  | **1/98 (1.0%)** | **1/55 (1.8%)** | **0/43 (0%)** |  |
| **MMSE, /30** | **92** | **27.0 (24.8, 28.0)** | **27.0 (25.2, 28.8)** | **26.0 (24.0, 28.0)** | **0.122** |
| **CDR® plus NACC FTLD-SB** | **84** | **3.0 (1.9, 5.0)** | **2.5 (1.5, 4.5)** | **4.2 (3.0, 5.5)** | **0.0032** |
| **Global Score CDR® plus NACC FTLD** | **84** |  |  |  | **0.135** |
| **0.5** |  | **40/84 (48%)** | **29/52 (56%)** | **11/32 (34%)** |  |
| **1** |  | **38/84 (45%)** | **19/52 (37%)** | **19/32 (59%)** |  |
| **2** |  | **6/84 (7.1%)** | **4/52 (7.7%)** | **2/32 (6.2%)** |  |
| **NPI total score** | **88** | **11.0 (3.8, 24.2)** | **10.0 (3.0, 21.5)** | **12.0 (6.0, 25.0)** | **0.52** |
| **Mutation** | **98** |  |  |  | **<0.0015** |
| **No Mutation** |  | **79/98 (81%)** | **40/55 (73%)** | **39/43 (91%)** |  |
| **Not Screened** |  | **16/98 (16%)** | **15/55 (27%)** | **1/43 (2.3%)** |  |
| **GRN** |  | **3/98 (3.1%)** | **0/55 (0%)** | **3/43 (7.0%)** |  |
| **Neuropathological Diagnosis** | **43** |  |  |  | **>0.95** |
| **Corticobasal Degeneration** |  | **17/43 (40%)** | **0/0 (NA%)** | **17/43 (40%)** |  |
| **Progressive Supranuclear Palsy** |  | **11/43 (26%)** | **0/0 (NA%)** | **11/43 (26%)** |  |
| **Pick's disease** |  | **7/43 (16%)** | **0/0 (NA%)** | **7/43 (16%)** |  |
| **FTLD-TDP type A** |  | **4/43 (9.3%)** | **0/0 (NA%)** | **4/43 (9.3%)** |  |
| **Other Pathologies** |  | **4/43 (9.3%)** | **0/0 (NA%)** | **4/43 (9.3%)** |  |

**Footnotes:** ^1^ Median (IQR); n (%); ^2^ Wilcoxon rank sum test; Pearson's Chi-squared test; Fisher's exact test; ^3^ False discovery rate correction for multiple testing

**Abbreviations:** CDR® plus NACC FTLD-SB = clinical dementia rating plus national Alzheimer’s coordinating center frontotemporal lobar degeneration sum of boxes; FTLD = frontotemporal lobar degeneration; MMSE = mini-mental state examination; NPI=neuropsychiatric inventory;

**Supplementary Table 3. Speech, language, and cognitive features of the participants with and without autopsy.**

| **Characteristic^1^** | **All Participants,**  **N = 102** | **No autopsy,**  **N = 59** | **Autopsy,**  **N = 43** | **p-value^2^** | **q-value^3^** |
| --- | --- | --- | --- | --- | --- |
| **Apraxia of Speech (MSE)** | 2.0 (1.0, 4.0) | 3.0 (2.0, 4.0) | 2.0 (1.0, 4.0) | 0.084 | 0.6 |
| **Dysarthria (MSE)** | 2.0 (0.0, 4.0) | 1.0 (0.0, 3.0) | 2.0 (0.8, 4.0) | 0.2 | 0.6 |
| **Syntax Comprehension (Bedside Screening)** | 4.0 (3.0, 5.0) | 5.0 (3.0, 5.0) | 4.0 (4.0, 4.0) | 0.4 | 0.7 |
| **Sequential Commands (WAB)** | 72.0 (67.0, 80.0) | 72.5 (67.0, 80.0) | 72.0 (68.5, 80.0) | 0.9 | >0.9 |
| **Mean Length of Utterance (SALT)** | 6.9 (4.4, 8.6) | 7.3 (4.9, 8.8) | 6.4 (4.0, 8.3) | 0.2 | 0.6 |
| **Subordinate Index (SALT)** | 1.0 (0.9, 1.1) | 1.0 (1.0, 1.1) | 1.0 (0.9, 1.1) | 0.5 | 0.8 |
| **Words per Minute (SALT)** | 54.4 (35.3, 76.8) | 57.6 (38.1, 77.2) | 50.9 (31.0, 69.5) | 0.4 | 0.7 |
| **Semantic Fluency (Animals, 1 min)** | 11.0 (7.0, 14.0) | 12.0 (8.0, 15.0) | 10.0 (6.0, 12.5) | 0.090 | 0.6 |
| **Confrontation Naming (BNT), /15** | 13.0 (11.0, 15.0) | 13.0 (11.0, 15.0) | 13.0 (11.8, 15.0) | 0.9 | >0.9 |
| **Single Word Comprehension (PPVT), /16** | 15.0 (13.0, 16.0) | 15.0 (13.0, 16.0) | 15.0 (14.0, 16.0) | 0.3 | 0.7 |
| **Repetition (WAB), /100** | 88.0 (75.8, 96.0) | 89.0 (79.2, 96.0) | 88.0 (72.0, 96.0) | 0.4 | 0.7 |
| **Verbal Working Memory (Digits backward)** | 3.0 (3.0, 4.0) | 4.0 (3.0, 4.0) | 3.0 (3.0, 4.0) | 0.090 | 0.6 |
| **Phonemic Fluency (D words, 1 min)** | 5.0 (3.0, 8.0) | 6.0 (3.0, 8.0) | 5.0 (3.0, 6.0) | 0.2 | 0.6 |
| **Stroop Test (Correct Words in the Inhibition Task in 1 min)** | 23.5 (15.8, 33.0) | 27.0 (19.0, 33.0) | 21.0 (14.0, 28.5) | 0.2 | 0.6 |
| **Set Shifting (Correct Modified Trails in 1 min)** | 14.0 (9.0, 14.0) | 14.0 (14.0, 14.0) | 14.0 (8.0, 14.0) | 0.2 | 0.6 |
| **Design Fluency (DKEFS)** | 6.0 (4.0, 8.0) | 6.0 (4.0, 8.0) | 6.0 (4.0, 8.0) | >0.9 | >0.9 |
| **Verbal Recognition (CVLT recognition), /9** | 9.0 (8.0, 9.0) | 9.0 (8.0, 9.0) | 8.0 (8.0, 9.0) | 0.4 | 0.7 |
| **Visual Memory (Benson figure), /17** | 11.0 (8.0, 13.0) | 12.0 (9.0, 13.0) | 10.0 (7.8, 12.0) | 0.2 | 0.6 |
| **Location discrimination (VOSP Number Location), /10** | 9.0 (8.0, 10.0) | 10.0 (8.0, 10.0) | 9.0 (8.0, 9.0) | 0.033 | 0.5 |
| **Visuoconstruction (Benson figure), /17** | 15.0 (13.5, 16.0) | 15.0 (14.0, 16.0) | 15.0 (13.0, 16.0) | 0.5 | 0.7 |
| **Calculations, /5** | 5.0 (4.0, 5.0) | 4.5 (4.0, 5.0) | 5.0 (4.0, 5.0) | 0.9 | >0.9 |
| **Hallucinations (NPI)** | 0 (0%) | 0 (0%) | 0 (0%) |  |  |
| **Delusions (NPI)** | 0 (0%) | 0 (0%) | 0 (0%) |  |  |
| **Dysphoria/Agression (NPI)** | 25 (29%) | 10 (20%) | 15 (43%) | 0.020 | 0.5 |
| **Depression (NPI)** | 30 (35%) | 17 (33%) | 13 (38%) | 0.6 | 0.8 |
| **Anxiety (NPI)** | 40 (47%) | 24 (46%) | 16 (47%) | >0.9 | >0.9 |
| **Euphoria/Elation (NPI)** | 8 (9.2%) | 3 (5.8%) | 5 (14%) | 0.3 | 0.7 |
| **Apathy/Indifference (NPI)** | 45 (52%) | 27 (52%) | 18 (53%) | >0.9 | >0.9 |
| **Disinhibition (NPI)** | 20 (24%) | 12 (24%) | 8 (24%) | >0.9 | >0.9 |
| **Irritability/Lability (NPI)** | 33 (39%) | 18 (35%) | 15 (44%) | 0.4 | 0.7 |
| **Aberrant Motor Behavior (NPI)** | 22 (26%) | 13 (25%) | 9 (26%) | 0.9 | >0.9 |
| **Sleep Changes (NPI)** | 24 (28%) | 13 (25%) | 11 (31%) | 0.5 | 0.8 |
| **NPI total score** | 10.0 (2.5, 21.0) | 9.5 (2.5, 20.2) | 12.0 (3.0, 23.5) | 0.5 | 0.7 |

**Footnotes:** ^1^ Median (IQR); n (%); ^2^ Wilcoxon rank sum test; Pearson's Chi-squared test; Fisher's exact test; ^3^ False discovery rate correction for multiple testing

**Abbreviations:** BNT=Boston Naming Test; DKEFS= Delis-Kaplan Executive Function Scale; MSE=Motor Speech Examination; NPI=neuropsychiatric inventory; PPVT= Peabody Picture Vocabulary Test; SALT= Systematic Analysis of Language Transcripts; VOSP= Visual Object Space Perception; WAB= Western Aphasia Battery.

**Supplementary Table 4. Frequency of main speech, language, and cognitive features.**

| **Characteristic** | **N** | **All Participants**, N = 98*^1^* | **AOS+agrammatism**, N = 69*^1^* | **PPAOS**, N = 18*^1^* | **Non-Fluent Dysarthric**, N = 7*^1^* | **PAA**, N = 4*^1^* | **p-value***^2^* |
| --- | --- | --- | --- | --- | --- | --- | --- |
| **Apraxia of Speech** | 98 | 87/98 (89%) | 69/69 (100%) | 18/18 (100%) | 0/7 (0%) | 0/4 (0%) | <0.001 |
| **Dysarthria** | 98 | 66/98 (67%) | 46/69 (67%) | 12/18 (67%) | 5/7 (71%) | 3/4 (75%) | >0.9 |
| **Motor Speech Impairment** | 98 | 95/98 (97%) | 69/69 (100%) | 18/18 (100%) | 5/7 (71%) | 3/4 (75%) | 0.001 |
| **Impaired Syntax Comprehension (Bedside Screening)** | 85 | 51/85 (60%) | 46/60 (77%) | 0/14 (0%) | 2/7 (29%) | 3/4 (75%) | <0.001 |
| **Impaired Syntax Comprehension (CYCLE)** | 87 | 48/87 (55%) | 43/62 (69%) | 0/15 (0%) | 3/7 (43%) | 2/3 (67%) | <0.001 |
| **Impaired Sequential Commands (WAB)** | 93 | 44/93 (47%) | 38/68 (56%) | 2/15 (13%) | 1/7 (14%) | 3/3 (100%) | <0.001 |
| **Reduced Mean Length of Utterance (SALT)** | 98 | 38/98 (39%) | 37/69 (54%) | 0/18 (0%) | 0/7 (0%) | 1/4 (25%) | <0.001 |
| **Reduced Subordination Index (SALT)** | 98 | 12/98 (12%) | 11/69 (16%) | 0/18 (0%) | 0/7 (0%) | 1/4 (25%) | 0.2 |
| **Morphosyntactic Errors (SALT)** | 98 | 45/98 (46%) | 41/69 (59%) | 0/18 (0%) | 0/7 (0%) | 4/4 (100%) | <0.001 |
| **Receptive Agrammatism** | 94 | 64/94 (68%) | 57/67 (85%) | 0/16 (0%) | 4/7 (57%) | 3/4 (75%) | <0.001 |
| **Expressive Agrammatism** | 98 | 57/98 (58%) | 53/69 (77%) | 0/18 (0%) | 0/7 (0%) | 4/4 (100%) | <0.001 |
| **Agrammatism (Expressive or Receptive)** | 98 | 77/98 (79%) | 69/69 (100%) | 0/18 (0%) | 4/7 (57%) | 4/4 (100%) | <0.001 |
| **Reduced Phonemic Fluency (D words, 1 min)** | 92 | 77/92 (84%) | 61/65 (94%) | 7/16 (44%) | 6/7 (86%) | 3/4 (75%) | <0.001 |
| **Reduced Words per Minute (SALT)** | 98 | 78/98 (80%) | 60/69 (87%) | 9/18 (50%) | 5/7 (71%) | 4/4 (100%) | 0.005 |
| **Impaired Confrontation Naming** | 93 | 45/93 (48%) | 38/66 (58%) | 1/16 (6.2%) | 4/7 (57%) | 2/4 (50%) | <0.001 |
| **Impaired Word Comprehension** | 87 | 42/87 (48%) | 34/61 (56%) | 3/15 (20%) | 2/7 (29%) | 3/4 (75%) | 0.031 |
| **Executive Impairment (Verbal Measures)** | 93 | 78/93 (84%) | 60/66 (91%) | 8/16 (50%) | 6/7 (86%) | 4/4 (100%) | 0.002 |
| **Executive Impairment (Non-Verbal Measures)** | 93 | 61/93 (66%) | 45/66 (68%) | 7/16 (44%) | 6/7 (86%) | 3/4 (75%) | 0.2 |
| **Memory Impairment** | 94 | 32/94 (34%) | 24/67 (36%) | 2/16 (12%) | 4/7 (57%) | 2/4 (50%) | 0.10 |
| **Visuospatial Impairment** | 94 | 36/94 (38%) | 28/67 (42%) | 5/16 (31%) | 2/7 (29%) | 1/4 (25%) | 0.8 |
| **Impaired Calculations** | 94 | 39/94 (41%) | 30/67 (45%) | 3/16 (19%) | 3/7 (43%) | 3/4 (75%) | 0.13 |
| **Any Behavioral Change** | 98 | 77/98 (88%) | 55/69 (87%) | 13/18 (81%) | 7/7 (100%) | 2/4 (100%) | 0.3 |

**Footnotes:** ^1^ n/N (%); ^2^ False discovery rate correction for multiple testing

**Abbreviations:** MSE=Motor Speech Examination; NPI=neuropsychiatric inventory; SALT= Systematic Analysis of Language Transcripts; WAB= Western Aphasia Battery.

**Supplementary Table 5. Comparison of clinical features between clinical subgroups**

| **Characteristic** | **N** | **All Participants**, N = 98*^1^* | **Clinical Subgroups** | | | | **p-value** | **q-value***^2^* |
| --- | --- | --- | --- | --- | --- | --- | --- | --- |
|  |  |  | **AOS+agrammatism**, N = 69*^1^* | **PPAOS**, N = 18*^1^* | **Non-Fluent Dysarthric**, N = 7*^1^* | **PAA**, N = 4*^1^* |  |  |
| **Syntax Comprehension (CYCLE)** | 87 | -4.8 (6.4) | -6.0 (6.4) | 0.1 (0.9) | -2.8 (5.9) | -11.1 (10.5) | <0.001*^3^* | <0.001 |
| **Syntax Comprehension (Bedside Screening)** | 85 | -1.5 (1.6) | -1.9 (1.5) | 0.0 (0.0) | -0.4 (0.7) | -2.3 (2.0) | <0.001*^3^* | <0.001 |
| **Sequential Commands (WAB)** | 93 | -1.3 (1.8) | -1.5 (1.8) | -0.2 (0.4) | -0.3 (0.5) | -3.1 (2.8) | 0.001*^3^* | 0.002 |
| **Mean Length of Utterance (SALT)** | 98 | -1.5 (1.3) | -1.9 (1.1) | -0.7 (0.7) | -1.1 (0.5) | 1.2 (3.1) | <0.001*^3^* | <0.001 |
| **Subordination Index (SALT)** | 98 | -1.2 (1.2) | -1.3 (1.3) | -0.6 (0.5) | -0.7 (0.5) | -1.9 (1.9) | 0.13*^3^* | 0.15 |
| **Morphosyntactic Errors, (SALT)** | 98 | -2.2 (2.8) | -3.0 (2.7) | 0.3 (0.9) | 0.1 (1.3) | -4.3 (2.3) | <0.001*^3^* | <0.001 |
| **Words per Minute (SALT)** | 98 | -3.3 (1.2) | -3.7 (1.1) | -2.0 (1.1) | -2.7 (0.9) | -3.3 (0.3) | <0.001*^3^* | <0.001 |
| **Semantic Fluency (Animals, 1 min)** | 91 | -2.2 (1.3) | -2.5 (1.0) | -0.8 (1.4) | -2.4 (1.7) | -3.0 (1.4) | <0.001*^3^* | 0.001 |
| **Confrontation Naming (BNT)** | 93 | -1.9 (3.0) | -2.3 (2.9) | 0.3 (0.7) | -2.1 (4.0) | -3.5 (4.2) | <0.001*^3^* | <0.001 |
| **Single Word Comprehension (PPVT)** | 87 | -2.4 (4.0) | -2.9 (4.0) | -0.5 (2.1) | -2.9 (6.6) | -2.4 (2.6) | 0.050*^3^* | 0.061 |
| **Repetition (WAB)** | 92 | -7.0 (7.4) | -8.3 (8.0) | -2.3 (2.4) | -3.6 (4.8) | -8.2 (4.8) | 0.002*^3^* | 0.003 |
| **Verbal Working Memory (Digits backward)** | 92 | -1.7 (1.2) | -1.9 (1.0) | -0.9 (1.3) | -1.2 (1.5) | -2.6 (1.1) | 0.019*^3^* | 0.028 |
| **Phonemic Fluency (D words, 1 min)** | 92 | -2.2 (0.9) | -2.4 (0.8) | -1.3 (1.3) | -2.3 (0.5) | -2.3 (0.8) | 0.013*^3^* | 0.023 |
| **Stroop Test (Correct Words in the Inhibition Task in 1 min)** | 75 | -2.7 (1.2) | -3.0 (1.1) | -1.6 (1.1) | -2.8 (0.8) | -3.6 (0.7) | 0.001*^3^* | 0.003 |
| **Set Shifting (Correct Modified Trails in 1 min)** | 89 | -1.9 (3.4) | -1.7 (3.2) | -0.7 (2.8) | -4.4 (4.4) | -4.8 (3.7) | 0.023*^3^* | 0.032 |
| **Design Fluency (DKEFS)** | 93 | -1.6 (1.0) | -1.7 (1.0) | -0.9 (1.0) | -1.9 (0.3) | -2.4 (1.2) | 0.016*^3^* | 0.026 |
| **Verbal Recognition (CVLT recognition)** | 89 | -0.5 (1.8) | -0.5 (1.7) | 0.0 (1.0) | -1.4 (3.5) | 0.6 (0.0) | 0.4*^3^* | 0.5 |
| **Visual Memory (Benson figure)** | 93 | -0.5 (1.3) | -0.6 (1.1) | 0.6 (0.5) | -1.0 (1.9) | -1.6 (2.7) | <0.001*^3^* | 0.001 |
| **Location discrimination (VOSP Number Location)** | 86 | -0.4 (1.3) | -0.3 (1.3) | -0.2 (1.2) | -0.9 (2.2) | -0.4 (1.5) | 0.9*^3^* | 0.9 |
| **Visuoconstruction (Benson figure)** | 91 | -0.9 (1.8) | -1.0 (1.7) | -0.2 (1.4) | -1.7 (2.8) | -0.3 (1.1) | 0.3*^3^* | 0.4 |
| **Calculations** | 94 | -1.3 (2.2) | -1.4 (2.3) | -0.2 (1.2) | -1.3 (1.9) | -2.5 (3.2) | 0.044*^3^* | 0.057 |

**Footnotes:** *^1^* Mean (SD); ^2^ False discovery rate correction for multiple testing; ^3^ Kruskal-Wallis rank sum test

**Abbreviations:** BNT=Boston Naming Test; DKEFS= Delis-Kaplan Executive Function Scale; MSE=Motor Speech Examination; PPVT= Peabody Picture Vocabulary Test; SALT= Systematic Analysis of Language Transcripts; VOSP= Visual Object Space Perception; WAB= Western Aphasia Battery.

**Supplementary Table 6. Comparison of clinical features between neuropathological groups**

| **Characteristic** | **N** | **All Participants,**  **N = 43*^1^*** | **Clinical Subgroups** | | | | | **p-value** | **q-value*^2^*** |
| --- | --- | --- | --- | --- | --- | --- | --- | --- | --- |
|  |  |  | **Progressive Supranuclear Palsy,**  **N = 11*^1^*** | **Corticobasal Degeneration,**  **N = 17*^1^*** | **Pick's disease, N = 7*^1^*** | **FTLD-TDP type A,**  **N = 4*^1^*** | **Other Pathologies,**  **N = 4*^1^*** |  |  |
| **Apraxia of Speech (MSE)** | 43 | 2.2 (1.9) | 1.6 (2.0) | 1.8 (1.1) | 3.4 (2.9) | 3.5 (1.7) |  | 0.3*^3^* | 0.8 |
| **Dysarthria (MSE)** | 43 | 2.0 (1.9) | 2.8 (1.5) | 1.5 (1.5) | 1.4 (1.8) | 0.2 (0.5) | 4.0 (2.7) | 0.022*^3^* | 0.4 |
| **Syntax Comprehension (CYCLE)** | 43 | 87.8 (13.7) | 96.1 (5.4) | 81.3 (16.6) | 90.4 (10.4) | 85.0 (13.7) | 91.2 (11.4) | 0.073*^3^* | 0.4 |
| **Syntax Comprehension (Bedside Screening)** | 35 | 4.0 (0.9) | 4.6 (0.5) | 3.8 (1.0) | 3.8 (0.8) | 4.2 (1.0) | 3.5 (0.6) | 0.14*^3^* | 0.6 |
| **Sequential Commands (WAB)** | 43 | 69.9 (13.4) | 78.8 (2.7) | 65.6 (15.5) | 66.9 (15.3) | 67.0 (16.7) | 71.8 (6.9) | 0.032*^3^* | 0.4 |
| **Mean Length of Utterance (SALT)** | 43 | 5.8 (3.2) | 6.8 (1.6) | 5.2 (2.1) | 6.4 (6.8) | 4.7 (1.8) | 5.5 (2.5) | 0.3*^3^* | 0.8 |
| **Subordination Index (SALT)** | 43 | 0.9 (0.4) | 1.0 (0.1) | 0.8 (0.4) | 0.6 (0.5) | 0.8 (0.6) | 0.9 (0.4) | 0.5*^3^* | 0.9 |
| **Morphosyntactic Errors, % (SALT)** | 43 | 0.5 (0.3) | 0.8 (0.2) | 0.6 (0.3) | 0.3 (0.4) | 0.5 (0.4) | 0.4 (0.3) | 0.046*^3^* | 0.4 |
| **Words per Minute (SALT)** | 42 | 54.7 (29.7) | 68.2 (28.9) | 50.3 (27.2) | 40.9 (31.7) | 68.6 (34.3) | 44.6 (28.5) | 0.3*^3^* | 0.8 |
| **Semantic Fluency (Animals, 1 min)** | 40 | 10.3 (6.0) | 11.8 (6.2) | 8.2 (3.9) | 12.0 (1.2) | 14.8 (12.8) | 8.5 (5.2) | 0.4*^3^* | 0.8 |
| **Confrontation Naming (BNT), /15** | 42 | 12.6 (2.7) | 13.1 (1.8) | 12.7 (3.0) | 10.8 (3.3) | 14.0 (1.4) | 12.0 (3.4) | 0.3*^3^* | 0.8 |
| **Single Word Comprehension (PPVT), /16** | 38 | 14.5 (2.3) | 14.7 (1.8) | 14.2 (2.8) | 15.2 (1.1) | 14.2 (2.4) | 14.0 (3.4) | >0.9*^3^* | >0.9 |
| **Repetition (WAB), /100** | 43 | 84.0 (18.4) | 91.3 (8.5) | 84.1 (18.4) | 72.7 (29.5) | 86.5 (10.0) | 81.2 (19.3) | 0.7*^3^* | >0.9 |
| **Verbal Working Memory (Digits backward)** | 41 | 3.1 (1.4) | 3.5 (1.8) | 3.0 (1.2) | 3.0 (0.9) | 3.2 (1.3) | 2.2 (1.7) | 0.7*^3^* | >0.9 |
| **Phonemic Fluency (D words, 1 min)** | 41 | 5.2 (4.1) | 5.8 (3.6) | 3.9 (2.0) | 5.8 (2.2) | 8.8 (9.8) | 5.0 (5.1) | 0.5*^3^* | 0.9 |
| **Stroop Test (Correct Words in the Inhibition Task in 1 min)** | 34 | 21.3 (11.2) | 22.2 (9.8) | 20.4 (12.1) | 24.4 (7.8) | 22.0 (22.1) | 16.0 (9.5) | 0.8*^3^* | >0.9 |
| **Set Shifting (Correct Modified Trails in 1 min)** | 41 | 11.4 (4.0) | 10.3 (5.1) | 11.8 (3.5) | 12.5 (3.7) | 11.8 (2.6) | 10.7 (5.8) | 0.8*^3^* | >0.9 |
| **Design Fluency (DKEFS)** | 43 | 5.9 (3.2) | 5.9 (3.0) | 6.1 (3.4) | 5.0 (2.8) | 7.5 (5.3) | 4.8 (2.1) | 0.9*^3^* | >0.9 |
| **Verbal Recognition (CVLT recognition), /9** | 40 | 8.3 (1.0) | 8.1 (1.3) | 8.3 (1.0) | 8.6 (0.5) | 8.8 (0.5) | 7.8 (1.0) | 0.5*^3^* | 0.9 |
| **Visual Memory (Benson figure), /17** | 43 | 10.5 (3.2) | 9.6 (4.0) | 11.1 (3.2) | 10.3 (3.5) | 10.8 (1.5) | 10.5 (2.4) | >0.9*^3^* | >0.9 |
| **Location discrimination (VOSP Number Location), /10** | 37 | 8.5 (1.3) | 8.0 (1.9) | 8.8 (1.3) | 8.7 (0.8) | 8.5 (0.6) | 9.0 (1.0) | 0.7*^3^* | >0.9 |
| **Visuoconstruction (Benson figure), /17** | 43 | 14.3 (2.1) | 13.7 (2.1) | 14.3 (2.1) | 15.1 (2.0) | 14.2 (2.2) | 14.2 (2.9) | 0.7*^3^* | >0.9 |
| **Calculations, /5** | 43 | 4.2 (1.0) | 4.5 (0.8) | 4.4 (0.8) | 4.1 (0.7) | 3.8 (1.9) | 3.2 (1.3) | 0.3*^3^* | 0.8 |

**Footnotes:** *^1^* Mean (SD); *^2^* False discovery rate correction for multiple testing; *^3^* Kruskal-Wallis rank sum test.

**Abbreviations:** BDAE= Boston Diagnostic Aphasia Examination; BNT=Boston Naming Test; DKEFS= Delis-Kaplan Executive Function Scale; MSE=Motor Speech Examination; PPVT= Peabody Picture Vocabulary Test; SALT= Systematic Analysis of Language Transcripts; VOSP= Visual Object Space Perception; WAB= Western Aphasia Battery.

**Supplementary Table 7. Linear Mixed Effects Model** **for the estimation of CDR-SB based on raw clinical data.**

|  | **Beta** | **SE** | **DF** | **t-value** | ***p*-value** | **95% CI** |
| --- | --- | --- | --- | --- | --- | --- |
| Intercept | 1.720 | 1.973 | 143 | 0.872 | 0.385 | -2.15 ; 5.59 |
| **Time (years from baseline)** | **1.226** | **0.097** | **143** | **12.690** | **0.000** | **1.04 ; 1.42** |
| Age at baseline | 0.004 | 0.029 | 82 | 0.125 | 0.901 | -0.05 ; 0.06 |
| Sex (Woman) | -0.363 | 0.441 | 82 | -0.823 | 0.413 | -1.23 ; 0.5 |
| Apraxia of Speech (MSE) | 0.294 | 0.243 | 82 | 1.207 | 0.231 | -0.18 ; 0.77 |
| Dysarthria (MSE) | 0.162 | 0.234 | 82 | 0.695 | 0.489 | -0.3 ; 0.62 |
| Words per Minute (SALT) | 0.008 | 0.315 | 82 | 0.026 | 0.979 | -0.61 ; 0.63 |
| Expressive Agrammatism | -0.479 | 0.291 | 82 | -1.645 | 0.104 | -1.05 ; 0.09 |
| Receptive Agrammatism | -0.071 | 0.288 | 82 | -0.248 | 0.805 | -0.64 ; 0.49 |
| Executive Function (Verbal) | -0.351 | 0.264 | 82 | -1.331 | 0.187 | -0.87 ; 0.17 |
| **Executive Function (Non-verbal)** | **-0.579** | **0.257** | **82** | **-2.253** | **0.027** | **-1.08 ; -0.08** |
| Time * Apraxia of Speech (MSE) | 0.032 | 0.098 | 143 | 0.325 | 0.746 | -0.16 ; 0.22 |
| **Time * Dysarthria (MSE)** | **-0.603** | **0.142** | **143** | **-4.234** | **0.000** | **-0.88 ; -0.32** |
| **Time * Words per Minute (SALT)** | **-0.366** | **0.147** | **143** | **-2.497** | **0.014** | **-0.08 ; -0.65** |
| **Time * Expressive Agrammatism** | **-0.335** | **0.129** | **143** | **-2.604** | **0.010** | **-0.59 ; -0.08** |
| **Time * Receptive Agrammatism** | **-0.418** | **0.176** | **143** | **-2.378** | **0.019** | **-0.76 ; -0.07** |
| Time * Executive Function (Verbal) | -0.150 | 0.115 | 143 | -1.302 | 0.195 | -0.38 ; 0.08 |
| Time * Executive Function (Non-verbal) | -0.092 | 0.141 | 143 | -0.653 | 0.515 | -0.37 ; 0.18 |

**Abbreviations:** MSE=Motor Speech Examination; PPVT= Peabody Picture Vocabulary Test; SALT= Systematic Analysis of Language Transcripts; VOSP= Visual Object Space Perception; SE=standard error; DF=degrees of freedom; CI=confidence interval

**Supplementary Table 8. Linear Mixed Effects Model** **for the estimation of CDR-SB based on latent clinical dimensions.**

| Parameter | Beta | SE | DF | t-value | *p*-value | 95% CI |
| --- | --- | --- | --- | --- | --- | --- |
| Intercept | 1.755 | 1.699 | 145 | 1.033 | .303 | -1.58 ; 5.09 |
| Time (years from baseline) | 1.173 | 0.096 | 145 | 12.196 | **<.001** | 0.98 ; 1.36 |
| Age at baseline | 0.003 | 0.025 | 83 | 0.112 | .911 | -0.05 ; 0.05 |
| Sex (Woman) | -0.369 | 0.369 | 83 | -1.002 | .319 | -1.09 ; 0.35 |
| D1: Disease Severity & Agrammatism | -0.640 | 0.103 | 83 | -6.233 | **<.001** | -0.84 ; -0.44 |
| D2: Prominent Apraxia of Speech | 0.591 | 0.151 | 83 | 3.912 | **<.001** | 0.29 ; 0.89 |
| D3: Prominent Dysarthria | -0.127 | 0.187 | 83 | -0.681 | 0.498 | -0.49 ; 0.24 |
| Time * D1 | -0.278 | 0.047 | 145 | -5.873 | **<.001** | -0.37 ; -0.19 |
| Time * D2 | -0.037 | 0.057 | 145 | -0.645 | 0.520 | -0.15 ; 0.07 |
| Time * D3 | -0.183 | 0.101 | 145 | -1.820 | 0.071 | -0.38 ; 0.01 |

**Abbreviations:** SE=standard error; DF=degrees of freedom; CI=confidence interval

**Supplementary Figures**

**Supplementary Figure. Neuropathological diagnoses along the spectrum of progressive non-fluent speech and language disorders.**

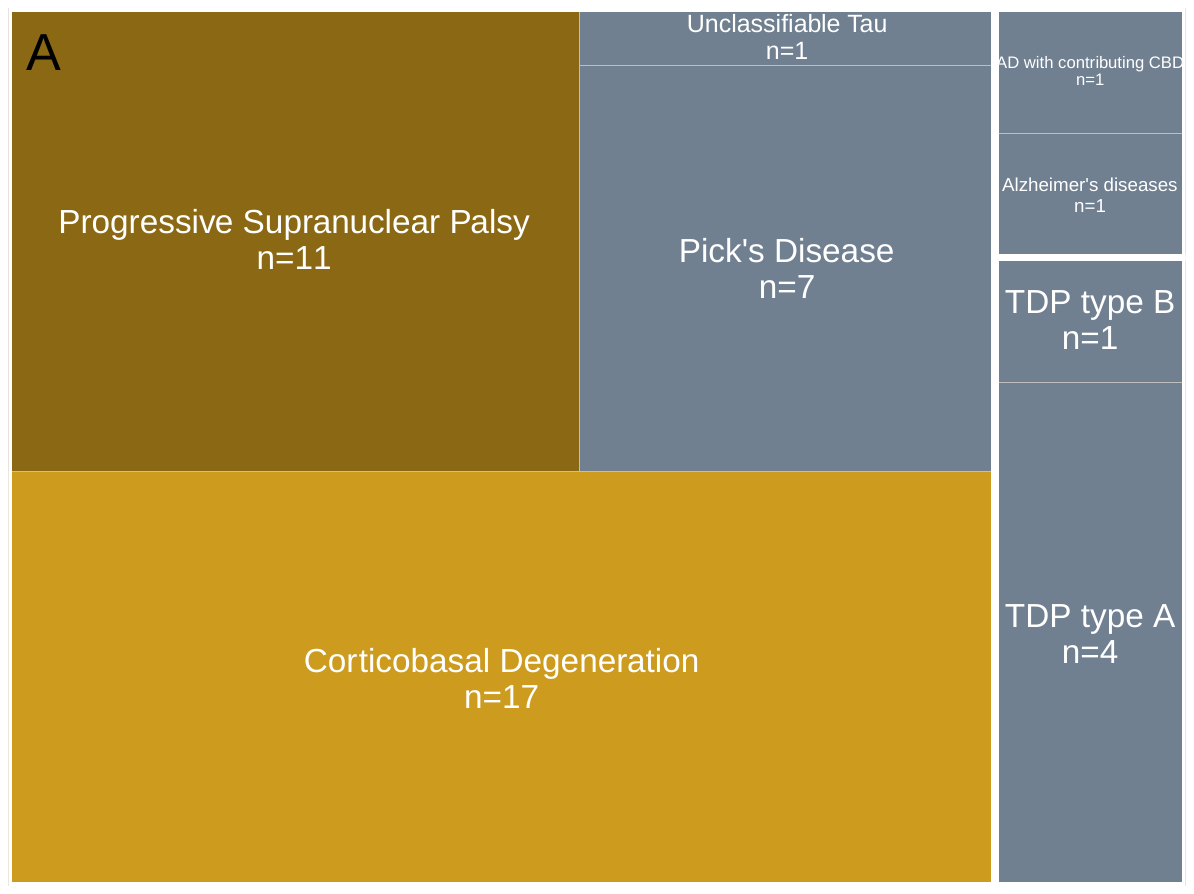

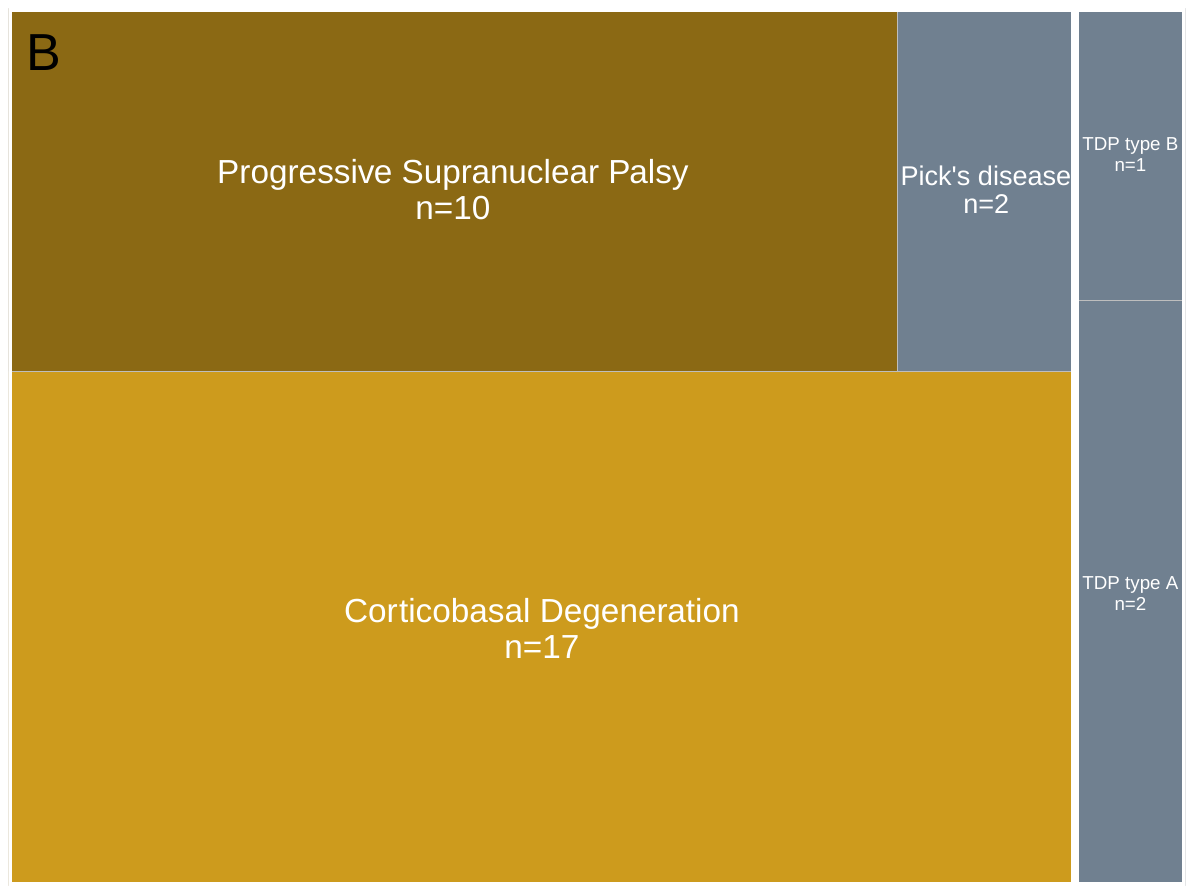

**Footnotes: A)** Neuropathological diagnoses along nf-SLD in the sample of this study; **B)** Neuropathological diagnosis along "progressive apraxia of speech" (Josephs et al., 2021).

**Abbreviations:** AD=Alzheimer's disease; CBD=Corticobasal degeneration.
